# Emulating the GRADE Trial Using Real-World Data

**DOI:** 10.1101/2022.05.23.22275392

**Authors:** Yihong Deng, Eric C. Polley, Joshua D. Wallach, Sanket S. Dhruva, Jeph Herrin, Kenneth Quinto, Charu Gandotra, William Crown, Peter Noseworthy, Xiaoxi Yao, Timothy D. Lyon, Nilay D. Shah, Joseph S. Ross, Rozalina G. McCoy

## Abstract

**OBJECTIVES:** To emulate the Glycemia Reduction Approaches in Diabetes: A Comparative Effectiveness Study (GRADE) trial using real-world data prior to its publication. GRADE is the first comparative effectiveness study to directly compare second-line glucose-lowering medications with respect to their ability to lower hemoglobin A_1c_ (HbA_1c_).

**DESIGN AND SETTING:** In this observational cohort study, we applied GRADE trial criteria to claims and laboratory data from OptumLabs^®^ Data Warehouse (OLDW), a U.S. nationwide claims database, between 1/25/2010 and 6/30/2019.

**PARTICIAPNTS:** Adults with type 2 diabetes with hemoglobin A_1c_ (HbA_1c_) 6.8-8.5% on metformin monotherapy, identified according to GRADE trial specifications.

**INTERVENTIONS:** Glimepiride, liraglutide, sitagliptin, and insulin glargine.

**MAIN OUTCOME MEASURES:** The primary outcome was time to HbA_1c_ ≥7.0% and secondary outcomes were other metabolic, microvascular, macrovascular, and safety outcomes specified by GRADE. Propensity scores were estimated using the gradient boosting machine method and inverse propensity score weighting was used to emulate randomization of the treatment groups, which were then compared using Cox proportional hazards regression.

**RESULTS:** We identified 8252 patients (19.7% of adults starting the study drugs in OLDW) meeting GRADE eligibility criteria (glimepiride arm=4318, liraglutide arm=690, sitagliptin arm=2993, glargine arm=251). The glargine arm was excluded from analyses due to small sample size. Median times to HbA_1c_ ≥7.0% were 442 (95% CI, 394-480) days for glimepiride, 764 (95% CI, 741-NA) days for liraglutide, and 427 (95% CI, 380-483) days for sitagliptin. Liraglutide was associated with lower risk of reaching HbA_1c_ ≥7.0% compared to glimepiride (HR 0.57 [95% CI, 0.43-0.75]) and sitagliptin (HR 0.55 [95% CI, 0.41-0.73]). Results were consistent for the secondary outcome of time to HbA_1c_ >7.5%. There were no significant differences among treatment groups for the remaining secondary outcomes.

**CONCLUSIONS:** In this emulation of the GRADE trial, liraglutide was significantly more effective at maintaining glycemic control than glimepiride or sitagliptin when added to metformin monotherapy. There is value in generating timely evidence on medical treatments using real-world data as a complement to prospective trials.

**SUMMARY BOXES:**

1. *What is already known about this topic?*
  - Real-world data is an important source of information about clinical practice, comparative effectiveness and safety, and health outcomes and has the potential to generate timely, pragmatic evidence on medical treatments as a complement to prospective clinical trials.
  - Multiple classes of second-line glucose-lowering medications have been approved for the management of type 2 diabetes, with limited evidence of their comparative effectiveness with respect to glycemic control.
2. *What this study adds?*
  - We emulated the Glycemia Reduction Approaches in Diabetes: A Comparative Effectiveness Study (GRADE) randomized clinical trial using data from a U.S. nationwide administrative claims database to identify the strengths and limitations of using real-world data to emulate prospective comparative effectiveness trials, particularly when examining medications in contexts that may not be the standard of care.
  - Liraglutide is more effective than glimepiride and sitagliptin at lowering HbA_1c_, supporting its preferential use when substantial glycemic reduction is needed.
  - Advanced causal inference analytic methods applied to observational data can be used to efficiently and effectively emulate clinical trials.

## INTRODUCTION

Type 2 diabetes is one of the most common serious chronic health conditions in the U.S. and worldwide, impacting 11.3% (37.3 million) of the U.S. population^1^ and 9.3% (463 million) people worldwide.^2^ Moderate glycemic control, defined by achieving glycosylated hemoglobin (HbA_1c_) between 7% and 8%, improves microvascular and macrovascular outcomes.^3 4^ Current clinical practice guidelines recommend targeting HbA_1c_ under 7% for most non-pregnant adults.^5^ Timely and appropriate treatment intensification is fundamental to maintaining glycemic control^6^ and preventing complications.^7-10^ Metformin is recommended as the first-line glucose-lowering medication due to its efficacy, tolerability, and low cost.^11-14^ However, type 2 diabetes is a progressive disease and most patients ultimately require intensification of therapy. Recent population-level estimates suggest that nearly one-third of patients with HbA_1c_ ≥7% are treated with only one glucose-lowering drug^15^ and as such would benefit from treatment intensification. Clinical practice guidelines advise that choice of second-line therapy should be informed by clinical and situational considerations specific to each patient, acknowledging the knowledge gaps stemming from the lack of direct comparisons of currently available second-line medications.^11-14^

The Glycemia Reduction Approaches in Diabetes: A Comparative Effectiveness Study (GRADE) is a recently completed, but still unpublished, pragmatic, randomized, parallel-arm clinical trial that seeks to address this knowledge gap by comparing four second-line glucose-lowering medications among adults with moderately uncontrolled type 2 diabetes on metformin monotherapy.^16 17^ The drugs represent four medication classes: glimepiride (sulfonylurea), sitagliptin (dipeptidyl-peptidase 4 inhibitor [DPP-4i]), liraglutide (glucagon-like peptide-1 receptor agonist [GLP-1RA]), and insulin glargine (basal analog insulin). GRADE was designed (2008) and launched (July 2013) prior to U.S. Food and Drug Administration (FDA) approval of sodium-glucose cotransporter-2 inhibitors (SGLT2i) and several cardiovascular outcomes trials that demonstrated reduction in atherosclerotic cardiovascular (ASCVD) and kidney disease outcomes with GLP-1RA use, and in heart failure and kidney disease outcomes with SGLT2i use. This highlights a key limitation of large prospective randomized controlled trials (RCTs): they are time-consuming to conduct, potentially hindering their ability to answer questions in a clinically meaningful time frame. Thus, there is value in efficiently generating timely evidence on medical treatments using observational research methods applied to real-world data as a complement to prospective trials.

Advances in the quantity, quality, and granularity of real-world data, combined with improvements in statistical methods used to account for confounding, treatment allocation bias, and time-related bias, have provided opportunities to use large-scale real-world data to inform our understanding of drug effectiveness and safety. Ideally, studies using real-world data would be conducted prior to the publication of RCT results, thereby minimizing potential biases that could be introduced by trying to replicate known RCT results. As an illustrative test case of the opportunities and limitations of using observational research methods to emulate RCTs, and building on parallel analyses emulating the PRONOUNCE trial,^18^ we used claims and laboratory results data from OptumLabs^®^ Data Warehouse (OLDW), a de-identified national dataset of privately insured and Medicare Advantage beneficiaries, to emulate the GRADE trial. We used published^16 19^ and publicly available^17^ information on GRADE’s study design to emulate the methods and anticipated results as closely as possible, with the goal of directly comparing the effectiveness of glimepiride, sitagliptin, liraglutide, and insulin glargine with respect to achieving and maintaining HbA_1c_ below 7.0% among adults with type 2 diabetes and HbA_1c_ 6.8-8.5% on metformin monotherapy. We also examined the secondary metabolic, microvascular, macrovascular, and safety endpoints planned in GRADE as feasible using the data available within OLDW. This work therefore has two complementary objectives. First – a clinical objective – to examine four second-line glucose-lowering medications with respect to lowering and/or maintaining HbA_1c_ below 7.0%, filling an important clinical knowledge gap with respect to the comparative effectiveness of these commonly used and guideline-recommended drug classes. Second – a methodologic objective – to ascertain whether routinely available claims data can be used to emulate a prospective randomized clinical trial ahead of its publication, filling important methodologic and regulatory policy needs with respect to the use of real-world data to predict clinical trial results.

## METHODS

### Study design

We retrospectively analyzed medical and pharmacy claims data from OLDW, a nationwide de-identified claims dataset comprised of enrollees in commercial and Medicare Advantage health plans. Private health plan beneficiaries are working-age adults and their dependents. Medicare Advantage plans are Medicare-approved plans offered by private companies to beneficiaries who are eligible for Medicare (e.g., adults 65 years and older, individuals with disability, patients with end-stage kidney disease) as a private alternative to Original Medicare. Just like commercial insurance, Medicare Advantage plans typically bundle medical and pharmacy coverage. OLDW contains longitudinal health information on enrollees in these health plans, representing a diverse mixture of ages, ethnicities, and geographic regions across the U.S.^20 21^ All study data are de-identified consistent with HIPAA expert de-identification determination. The study was exempt from Mayo Clinic Institutional Review Board review and is reported according to the Reporting of studies Conducted using Observational Routinely-collected Data (RECORD) reporting guideline.^22^

### Data Sharing Statement

OLDW data are available for research through a virtual data warehouse. The authors are not able to distribute the data. Study protocol, code sets, and statistical analysis plan are available online.^23^

### Study population

We first assembled a cohort of adults (≥18 years) who first started glimepiride, sitagliptin, liraglutide, or insulin glargine between January 25, 2010 (date of liraglutide approval by the FDA; remaining study drugs were approved earlier) and June 30, 2019 (**Figure S1 in the Supplement**). The index date was set to the date of the first claim for the study drug. Patients who started ≥2 study drugs on the index date were excluded. Patients were required to be adherent to metformin for ≥8 weeks prior to that first fill date. This was established by identifying all metformin fills prior to the index date, establishing continuous treatment episodes based on prescription fill dates and the days’ supply for each fill (allowing up to 30-day gap between fills), and requiring that the last metformin treatment episode prior to the index date be at least 8 weeks. To ensure consistent and adequate capture of baseline comorbidities and treatment data, patients were required to have 6 months of continuous enrollment with medical and pharmacy coverage before the index date. Patients with fills for any glucose-lowering medications other than metformin during the baseline period were excluded. Patients with type 1 diabetes, defined using International Classification of Diseases (ICD) codes were excluded. Patients were further required to have valid demographic (age, sex, region) data and HbA_1c_ results both within 3 months prior to the index date (“baseline HbA_1c_”) and during follow-up. Next, eligibility criteria for the GRADE trial^16 17 19^ were adapted and applied to beneficiaries included in OLDW as detailed in **Table S1**. All relevant diagnosis codes and medications are summarized in **Tables S2-S3**. All eligible patients in OLDW were included in the cohort.

### Outcomes

The primary outcome was time to primary metabolic failure of the assigned treatment, calculated as days to HbA_1c_ ≥7.0% while treated with the assigned medication, with the period of eligibility starting at month 3 after the index date (analogous to the first quarterly HbA_1c_ assessment in GRADE). Deviating from the GRADE protocol, we did not require a confirmatory HbA_1c_ due to variation in real-world HbA_1c_ testing intervals. To assess for potential bias in outcome ascertainment as the result of different frequencies of HbA_1c_ testing and varying intervals between tests among the treatment groups, we compared these number, frequency, and timing of available HbA_1c_ test results and found no difference between the groups (**Table S4**). Because testing frequency is guided by baseline HbA_1c_, we also examined intervals between sequential HbA_1c_ tests stratified by baseline HbA_1c_, and found no differences between the treatment groups (**Table S5**).

Secondary metabolic, cardiovascular, and microvascular outcomes were analyzed as specified in the GRADE statistical analysis plan (SAP)^17^ if they were feasible to ascertain using claims data (**Table S6)**. Patients were followed until they experienced the outcome of interest, anticipated follow-up duration of the trial (7 years), end of the study period (July 31, 2019), end of insurance coverage, or death. For outcomes observed “while being treated assigned regimen,” we followed patients until they discontinued the assigned medication (defined as not refilling a medication after 30 days of the end of last treatment episode), with the goal of emulating the definitions of these outcomes in the GRADE trial (i.e., “while treated with originally assigned medications”)^16^.

### Independent Variables

Patient individual-level age, sex, race/ethnicity, and annual household income were identified from OLDW enrollment files at the time of the index date. Detailed description of the source data for these variables is available in the **Supplemental Methods**. Comorbidities (ascertained from all claims during 6 months preceding the index date) included retinopathy, nephropathy, neuropathy, coronary artery disease, cerebrovascular disease, peripheral vascular disease, heart failure, and prior severe hypoglycemia and hyperglycemia as detailed in **Table S2**. Specialties of treating physicians were categorized as primary care, endocrinology, cardiology, nephrology, other, and unknown. Baseline medications, included as surrogates for complications burden, were identified from fills in the six months preceding the index date (**Table S3**).

### Patient and Public Involvement

Patients were not involved in the design, conduct, or dissemination of this study. However, this study was informed by the need to 1) identify preferred glucose-lowering treatment strategies in the absence of direct comparisons across the examined drugs; and 2) examine whether and how data collected in the process of routine patient care can be used to emulate prospective clinical trials. Because this study seeks to inform drug regulatory policy and procedures, investigators from the U.S. FDA contributed to the design of the study and interpretation of study findings; they are included as co-authors on this publication.

### Statistical analysis

Inverse probability of treatment weighting (IPTW) were used to balance the differences in baseline characteristics among the treatment groups. Propensity score (PS) were used as probability of treatment; these PS weights were estimated using Generalized Boosted Models (GBM) including the baseline variables presented in **Table 1**. GBM involves an iterative process with multiple regression trees to capture complex and nonlinear relationships between treatment assignments and the pretreatment covariates, resulting in the PS model leading to the best balance among the treatment groups.^24^ Additional detail on the models is provided in Supp. Methods. Stabilized weights with multiple treatments were calculated by dividing the marginal probability of treatment by the PS of treatment received.^25^ The distribution of weights is illustrated in **Figure S2**. Standardized mean differences (SMD) were used to assess the balance of covariates after weighting; SMD ≤0.1 was considered a good balance and SMD≤0.2 was considered acceptable.^26^ Prior to evaluation of the outcomes, weighted sample sizes and ability to account for baseline confounding were examined to determine the feasibility of including each treatment group.

**Table 1.** Baseline characteristics in weighted cohort. Data are presented as count (percentage), unless otherwise specified. ARB, angiotensin II receptor blockers; ACE, angiotensin-converting enzyme; HbA_1c_, hemoglobin A_1c_; SMD, standardized mean difference.

|  | **Glimepiride** | **Liraglutide** | **Sitagliptin** | **Largest SMD** |
| --- | --- | --- | --- | --- |
| **Weighted N** | 4168 | 572 | 2800 |  |
| **Age, years, mean (SD)** | 62.0 (11.1) | 60.5 (10.4) | 61.98 (11.0) | 0.14 |
| **Sex** |  |  |  | 0.05 |
| Female | 2009 (48.2%) | 289 (50.5%) | 1374 (49.1%) |  |
| Male | 2159 (51.8%) | 283 (49.5%) | 1427 (50.9%) |  |
| **Race/ethnicity** |  |  |  | 0.09 |
| White | 2695 (64.7%) | 376 (65.8%) | 1798 (64.2%) |  |
| Black | 536 (12.9%) | 79 (13.7%) | 355 (12.7%) |  |
| Hispanic | 513 (12.3%) | 72 (12.6%) | 353 (12.6%) |  |
| Asian | 266 (6.4%) | 28 (4.9%) | 185 (6.6%) |  |
| Unknown | 159 (3.8%) | 17 (3.0%) | 108 (3.9%) |  |
| **Annual Household Income** |  |  |  | 0.10 |
| <\$40,000 | 989 (23.7%) | 123 (21.6%) | 647 (23.1%) | |
| \$40,000 - \$74,999 | 1145 (27.5%) | 175 (30.7%) | 766 (27.3%) | |
| \$75,000 – \$124,999 | 1153 (27.7%) | 164 (28.6%) | 772 (27.6%) | |
| \$125,000 – \$199,999 | 485 (11.6%) | 63 (11.0%) | 336 (12.0%) | |
| ≥\$200,000 | 185 (4.4%) | 26 (4.5%) | 139 (4.9%) | |
| Unknown/missing | 211 (5.1%) | 21 (3.7%) | 142 (5.1%) |  |
| **Baseline HbA<sub>1c</sub>, mean (SD)** | 7.6 (0.5) | 7.6 (0.5) | 7.6 (0.5) | 0.06 |
| **Baseline HbA<sub>1c</sub> categories** |  |  |  | 0.09 |
| 6.8-6.9% | 336 (8.1%) | 59 (10.3%) | 233 (8.3%) |  |
| 7.0-7.9% | 2630 (63.1%) | 364 (63.7%) | 1786 (63.8%) |  |
| 8.0-8.5% | 1202 (28.8%) | 149 (26.0%) | 782 (27.9%) |  |
| **Baseline creatinine, mg/dL, mean (SD)\*** | 0.9 (0.2) | 0.9 (0.2) | 0.9 (0.2) | 0.02 |
| **Baseline comorbidities** |  |  |  |  |
| Nephropathy | 363 (8.7%) | 55 (9.5%) | 229 (8.2%) | 0.05 |
| Retinopathy | 196 (4.7%) | 31 (5.4%) | 135 (4.8%) | 0.03 |
| Neuropathy | 505 (12.1%) | 75 (13.2%) | 326 (11.7%) | 0.05 |
| Hyperglycemia | 2 (0.0%) | 0 | 0 | 0.03 |
| Hypoglycemia | 2 (0.0%) | 0 | 0 | 0.03 |
| Coronary artery disease | 355 (8.5%) | 53 (9.2%) | 224 (8.0%) | 0.04 |
| Chronic kidney disease | 121 (2.9%) | 18 (3.1%) | 82 (2.9%) | 0.01 |
| Cerebrovascular disease | 117 (2.8%) | 16 (2.8%) | 80 (2.8%) | 0.00 |
| Peripheral vascular disease | 190 (4.6%) | 28 (4.9%) | 130 (4.6%) | 0.02 |
| **Baseline medications** |  |  |  |  |
| ACE inhibitor | 1819 (43.6%) | 250 (43.7%) | 1206 (43.1%) | 0.01 |
| ARB | 1110 (26.6%) | 157 (27.4%) | 755 (27.0%) | 0.02 |
| ACE inhibitor or ARB | 2850 (68.4%) | 400 (69.9%) | 1906 (68.0%) | 0.04 |
| Direct oral anticoagulant | 57 (1.4%) | 9 (1.5%) | 40 (1.4%) | 0.01 |
| Statin | 2853 (68.5%) | 374 (65.4%) | 1937 (69.2%) | 0.08 |
| Non-statin lipid lowering medications | 549 (13.2%) | 85 (14.9%) | 382 (13.6%) | 0.05 |
| Warfarin | 83 (2.0%) | 12 (2.0%) | 48 (1.7%) | 0.02 |
| Peripheral neuropathy medications | 410 (9.8%) | 74 (12.9%) | 281 (10.0%) | 0.09 |
| **Specialty of treating physicians** |  |  |  | 0.10 |
| Primary care | 3202 (76.8%) | 424 (74.1%) | 2145 (76.6%) |  |
| Endocrinology | 174 (4.2%) | 27 (4.7%) | 120 (4.3%) |  |
| Cardiology | 28 (0.7%) | 2 (0.4%) | 19 (0.7%) |  |
| Nephrology | 6 (0.1%) | 0 | 5 (0.2%) |  |
| Other | 274 (6.6%) | 45 (7.9%) | 179 (6.4%) |  |
| Unknown | 483 (11.6%) | 74 (12.9%) | 333 (11.9%) |  |
| **Year of cohort entry** |  |  |  | 0.13 |
| 2010 | 237 (5.7%) | 29 (5.0%) | 167 (6.0%) |  |
| 2011 | 241 (5.8%) | 25 (4.4%) | 168 (6.0%) |  |
| 2012 | 297 (7.1%) | 41 (7.2%) | 204 (7.3%) |  |
| 2013 | 388 (9.3%) | 65 (11.3%) | 268 (9.6%) |  |
| 2014 | 464 (11.1%) | 65 (11.4%) | 318 (11.4%) |  |
| 2015 | 449 (10.8%) | 58 (10.1%) | 293 (10.4%) |  |
| 2016 | 575 (13.8%) | 75 (13.0%) | 380 (13.6%) |  |
| 2017 | 761 (18.3%) | 97 (16.9%) | 497 (17.8%) |  |
| 2018 | 632 (15.2%) | 91 (15.9%) | 422 (15.1%) |  |
| 2019 | 126 (3.0%) | 27 (4.7%) | 84 (3.0%) |  |

\*Baseline creatinine was not included in the propensity score model as there were missing values.

The cumulative incidence of the primary (time to first HbA_1c_ ≥7.0) and secondary (time to first HbA_1c_ >7.5%) metabolic failures within each treatment arm was estimated with the IPTW Kaplan-Meier method. PS weighted Cox proportional hazards (PH) regression models adjusted by baseline HbA_1c_ values were used to compare the outcomes between treatment groups. The at-risk time for the PH model was set as 3 months after the index date as the primary outcome can be only observed starting at 3^rd^ month. Results were presented as median times to metabolic failure and expected proportions of patients experiencing metabolic failure at 1 and 2 years. All pairwise comparisons between the treatment groups were estimated and the Holm method was applied to adjust the p-values for multiple testing. The proportional hazards assumption was tested using Schoenfeld residuals. Similar analyses were performed for other time-to-event outcomes. The at-risk start time for modeling secondary metabolic, cardiovascular, and microvascular disease outcomes was set at the study index date. Repeated measures HbA_1c_ trends by treatment group were estimated by using the IPTW mean HbA_1c_ results by treatment group in 3-month time intervals. The follow-up time by treatment arm was estimated using the same propensity score weights as the primary analysis and the IPTW Kaplan-Meier method for the censoring distribution.^27^

All primary analyses were conducted using the “per-protocol” censoring approach for the primary outcome and for the secondary outcomes of secondary metabolic failure and insulin initiation, censoring at the time of treatment drug discontinuation, disenrollment from the health plan, end of study period, or death, whichever came first (**Figure S3**). Time “on treatment” for each drug was determined by calculating continuous coverage episodes based on available fills, same as for baseline metformin therapy. Remaining secondary outcomes were analyzed using the “intention-to-treat” censoring approach, censoring the patient at the time of health plan disenrollment, end of study, or death, which ever came first. *P* < 0.05 was considered statistically significant for all 2-sided tests. All analyses were performed using SAS 9.4 (SAS Institute Inc., Cary, NC) and R version 4.0.2.(R Foundation).

### Subgroup analyses

A priori-defined subgroup analyses were performed as a function of baseline HbA_1c_ (<7.0% vs. ≥7.0%), age group (<65 years, ≥65 years), sex (male vs. female) and race/ethnicity (White, Black, Hispanic, Asian).

### Sensitivity analyses

First, to examine the comparative effectiveness of study medications while treated only with them and not with any other medication, accounting for real-world treatment practices, we repeated all analyses using the “as-treated” censoring approach, censoring at the time a new medication class was added, the assigned medication was discontinued, health plan disenrollment, end of study, or death, which ever came first (**Figure S3**). Second, we assessed residual confounding by testing a falsification end point that is unlikely to be associated with the studied medications: diagnosis of pneumonia (**Table S2**) during the follow-up period.

## RESULTS

### Study Population

We identified 18,365 adults with type 2 diabetes who started glimepiride, 12,818 who started sitagliptin, 5,021 who started liraglutide, and 5,659 who started insulin glargine and had the required baseline enrollment and available HbA_1c_ results (**Figure S1**). GRADE eligibility criteria were met by 19.7% of these patients, ranging from 4.4% of glargine-treated patients to 23.5% of glimepiride-treated patients. The most prevalent reasons for ineligibility (**Table S7**) were HbA_1c_ outside the prespecified range (ranging from 51.7% of sitagliptin-treated patients to 81.1% of glargine-treated patients) and not being treated with metformin monotherapy at the time of study drug initiation (ranging from 43.5% of glimepiride-treated patients to 68.5% of glargine-treated patients). The final cohort was comprised of 4318 patients in the glimepiride arm, 2993 patients in the sitagliptin arm, 690 patients in the liraglutide arm, and 251 patients in the glargine arm (see **Table S8** for all included drugs).

Baseline patient characteristics prior to weighting are shown in **Table S9**. There were significant differences (largest SMD >0.2) in patient age, race/ethnicity, annual household income, and prescribing physician specialty across the four treatment groups. Patients in the liraglutide arm were more likely to be younger, White, higher income, and treated by an endocrinologist than patients in the other treatment arms. Patients in the glargine arm were most likely to be low income and had the highest prevalence of all examined comorbidities.

The glargine arm was excluded from all analyses because of small sample size (N=251, weighted N=179) and inability to achieve good confounder control after weighting. After the glargine group was dropped, the PS model was estimated on the remaining 3 groups. After weighting, mean patient ages were 62.0 (SD, 11.1) years in the glimepiride arm, 62.0 (SD, 11.0) in the sitagliptin arm, and 60.5 (SD, 10.4) in the liraglutide arm (**Table 1**). Women comprised 48.2%, 49.1%, and 50.5% of the treatment arms, respectively. White patients comprised 64.7%, 64.2%, and 65.8% of the treatment arms, respectively. Mean baseline HbA_1c_ levels were 7.63% (SD, 0.48), 7.61% (SD, 0.47), and 7.60% (SD, 0.48), respectively. Pairwise SMDs for all baseline covariates are presented in **Table S10**; all SMDs were <0.2.

### Primary Metabolic Failure (HbA_1c_ ≥7.0%)

Median follow-up until per-protocol censoring was 238 (95% CI, 226-255) days in the glimepiride arm, 124 (95% CI, 100-150) days in the liraglutide arm, and 186 (95% CI, 179-201) days in the sitagliptin arm (**Figure S4**). Mean HbA_1c_ decreased most in the liraglutide arm and least in the sitagliptin arm, with differences most pronounced between months 3 and 6 of treatment (**Figure 1A**). The median times to primary metabolic failure were 442 (95% CI, 394-480) days in the glimepiride arm, 764 (95% CI, 741-NA) days in the liraglutide arm, and 427 (95% CI, 380-483) days in the sitagliptin arm (**Figure 1B**). Liraglutide was associated with lower risk of primary metabolic failure compared to glimepiride (HR 0.57; 95% CI, 0.43-0.75) and sitagliptin (HR 0.55; 95% CI, 0.41-0.73); **Table 2**. No significant difference was observed between sitagliptin and glimepiride (HR 1.03; 95% CI, 0.94-1.13). By 1-year, the estimated rates of primary metabolic failure were 0.28 (95% CI, 0.19-0.36) in the liraglutide arm, 0.44 (95% CI, 0.42-0.46) in the glimepiride arm, and 0.46 (95% CI, 0.43-0.48) in the sitagliptin arm (**Table 3**). The difference in event rates persisted at 2 years.

**Table 2.** Hazard ratios for primary and secondary metabolic outcomes. *Holm-adjusted p-values.

|  | **Hazard Ratio (95% CI)** | **P-value \*** |
| --- | --- | --- |
| **Primary metabolic failure (HbA<sub>1c</sub> ≥7%)** |  |  |
| Liraglutide vs Glimepiride | 0.57 (0.43, 0.75) | <0.001 |
| Sitagliptin vs Glimepiride | 1.03 (0.94, 1.13) | 0.48 |
| Liraglutide vs Sitagliptin | 0.55 (0.41, 0.73) | <0.001 |
| **Secondary metabolic failure (HbA<sub>1c</sub> >7.5%)** |  |  |
| Liraglutide vs Glimepiride | 0.61 (0.43, 0.87) | 0.01 |
| Sitagliptin vs Glimepiride | 1.04 (0.91, 1.18) | 0.60 |
| Liraglutide vs Sitagliptin | 0.59 (0.41, 0.85) | 0.01 |
| **Initiating insulin** |  |  |
| Liraglutide vs Glimepiride | 1.49 (0.76, 2.93) | 0.75 |
| Sitagliptin vs Glimepiride | 1.10 (0.77, 1.58) | 0.81 |
| Liraglutide vs Sitagliptin | 1.35 (0.67, 2.72) | 0.81 |

**Table 3.** Rates of primary and secondary metabolic failure by treatment arm.

|  | 1-year Event Rate (95% CI) | 2-year Event Rate (95% CI) |
| --- | --- | --- |
| **Primary metabolic failure ($\text{HbA}_{1c} \geq 7\%$)** |  |  |
| Glimepiride | 0.44 (0.42, 0.46) | 0.65 (0.63, 0.68) |
| Sitagliptin | 0.28 (0.19, 0.36) | 0.40 (0.29, 0.49) |
| Liraglutide | 0.46 (0.43, 0.48) | 0.62 (0.58, 0.65) |
| **Secondary metabolic failure ($\text{HbA}_{1c} > 7.5\%$)** |  |  |
| Glimepiride | 0.20 (0.19, 0.22) | 0.37 (0.34, 0.39) |
| Sitagliptin | 0.11 (0.06, 0.17) | 0.17 (0.10, 0.25) |
| Liraglutide | 0.22 (0.19, 0.24) | 0.37 (0.33, 0.41) |

**Figure 1.**
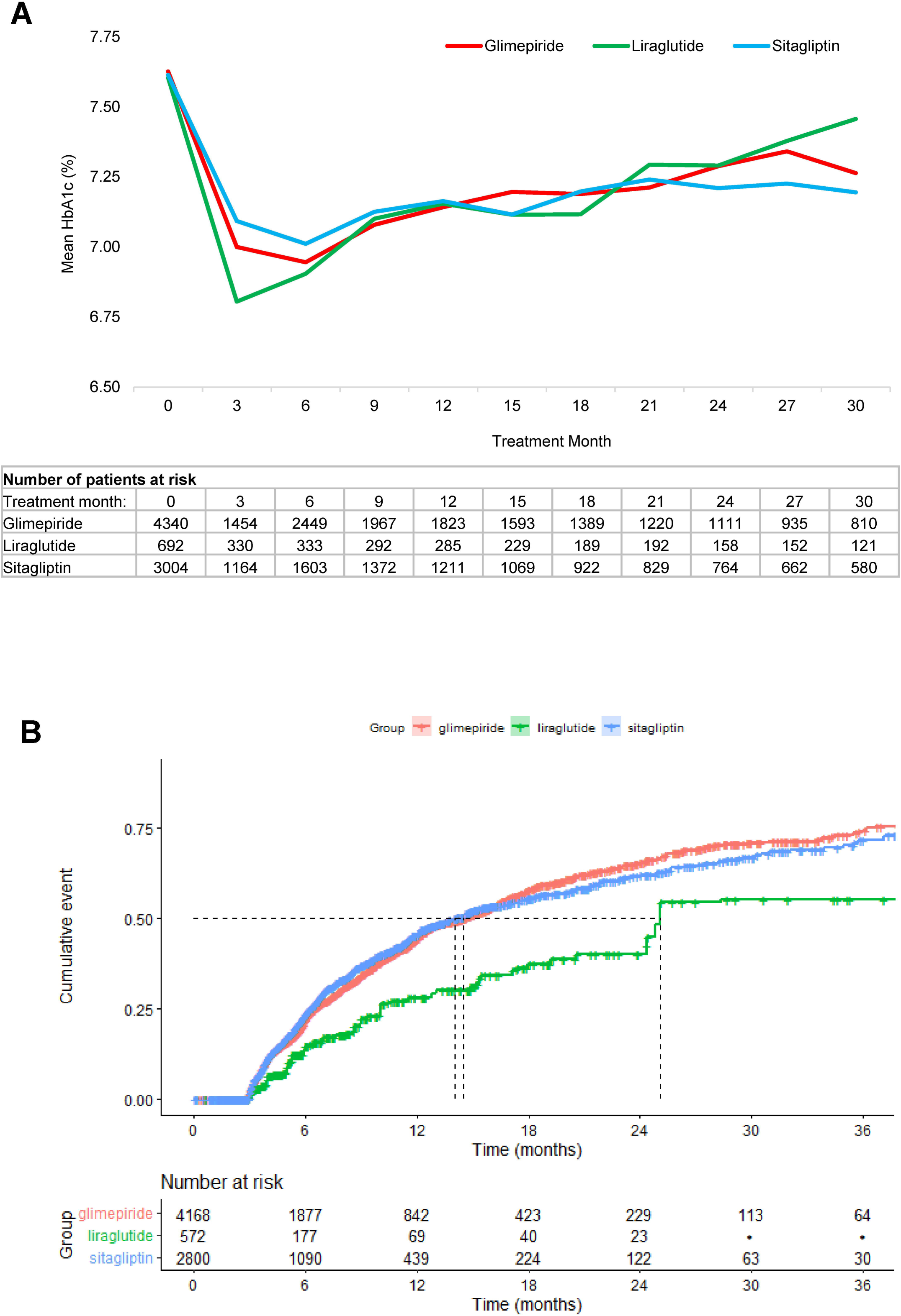
Glycemic control. (A) Mean HbA_1c_ levels over time. Results are based on observed on-treatment trajectories, with no imputation of missing HbA_1c_ levels. (B) Cumulative risks of primary metabolic failure in propensity score weighted patients.

### Secondary Metabolic Failure (HbA_1c_ >7.5%)

Time to secondary metabolic failure was longest in the liraglutide arm (**Figure S5**). Liraglutide was associated with lower risk of secondary metabolic failure compared to glimepiride (HR 0.61; 95% CI [0.43, 0.87]) and sitagliptin (HR 0.59; 95% CI [0.41, 0.85]); (**Table 2)**. By 1-year, the estimated rates of secondary metabolic failure were 0.11 (95% CI, 0.06-0.17) in the liraglutide arm, 0.20 (95% CI, 0.19-0.22) in the glimepiride arm, and 0.22 (95% CI, 0.19-0.24) in the sitagliptin arm (**Table 3**). The difference in event rates persisted at 2 years.

### Other Secondary Outcomes

Insulin was started by 84 (2.0%) patients in the glimepiride arm, 11 (1.9%) patients in the liraglutide arm, and 50 (1.8%) patients in the sitagliptin arm, with no significant difference among the three groups **(Table 2)**. Overall, 37 patients experienced emergency department visits or hospitalizations for hypoglycemia during the study period, including <11 in the liraglutide and sitagliptin arms, precluding statistical analyses.

Heart failure, ESKD, pancreatitis, pancreatic/thyroid cancer, and all-cause mortality could not be analyzed due to <11 events in all treatment groups (event rates are presented in **Table S11**). There were no statistically significant differences between groups for MACE, retinopathy, neuropathy, other cardiovascular events, cancer, and all-cause hospitalizations **(Table S12)**.

### Subgroup Analyses

Liraglutide was associated with lower risk of primary metabolic failure compared with glimepiride (HR 0.59; 95% CI, 0.44-0.78) and sitagliptin (HR 0.58; 95% CI, 0.43-0.79) among patients with baseline HbA_1c_ ≥7.0%. There were no significant differences among the treatment groups in patients with baseline HbA_1c_ <7.0% (**Table S13**). Liraglutide was associated with lower risk of primary metabolic failure compared with glimepiride (HR 0.54; 95% CI, 0.42-0.71) and sitagliptin (HR 0.58; 95% CI, 0.44-0.77) among patients <65 years old. There were no significant differences among groups in patients ≥65 years of age. Liraglutide was also associated with lower risks of primary metabolic failure than glimepiride and sitagliptin in women, but not in men, and in White and Hispanic patients, but not in Black or Asian patients. Findings were similar for secondary metabolic failure **(Table S14**).

### Sensitivity Analyses

Another glucose-lowering medication was added prior to discontinuation of the assigned treatment in 434 (10%) patients in the glimepiride arm, 268 (39%) in the liraglutide arm, and 429 (14%) in the sitagliptin arm. Sensitivity analyses using the as-treated censor approach were consistent with the primary analyses (**Figure S6, Table S15**). There were no significant differences among the treatment groups in the pneumonia falsification endpoint **(Table S16)**.

## DISCUSSION

Our emulation of the GRADE trial using real-world data from an administrative claims database revealed that liraglutide was significantly more effective at maintaining glycemic control, defined by time to HbA_1c_ ≥7.0% (primary metabolic failure) and HbA_1c_ >7.5% (secondary metabolic failure) than either glimepiride or sitagliptin. These differences are clinically meaningful, with over 40% more patients in control of their HbA_1c_ when treated with liraglutide than when treated with glimepiride or sitagliptin. We were unable to include insulin glargine in the comparisons because of the small number of glargine-treated patients meeting GRADE eligibility criteria. This was not surprising as treatment with basal insulin in the clinical context examined by GRADE is outside the standard of care and mainstream practice. Additionally, the analytic framework implemented in this work demonstrates that real-world data may be an important complement to prospective trials, allowing for efficient and timely examination of pressing clinical questions and inquiries of comparative effectiveness and safety.

Our efforts to emulate all specifications of the GRADE trial were hindered by the fact that study conditions are not adequately represented in real-world practice as they are not supported by clinical practice guidelines. While all four study drugs were frequently used by the OLDW population, 80% of adults starting these medications had to be excluded because they did not meet the prespecified GRADE eligibility criteria. Nevertheless, this proportion of included patients is still higher than the 9.1% generalizability estimated by the GRADE study team when compared to the overall U.S. population with diabetes.^19^ The majority of patients (58.6% overall) were excluded due to not meeting baseline HbA_1c_ level requirements, including 81.1% of glargine-initiators, 71.8% of liraglutide-initiators, 52.8% of glimepiride initiators, and 51.7% of sitagliptin-initiators. According to current guidelines, the target HbA_1c_ for most non-pregnant adults is 7.0%, such that treatment intensification would not be warranted for some participants. Initiation of insulin, in particular, is advised when HbA_1c_ is above 9-10%,^14 28^ so starting glargine as a second-line drug at HbA_1c_ levels below 8.5% would not be consistent with the standard of care^14 28^ or contemporary practice.^29-31^ The fact that most patients treated with the examined medications in clinical practice are not represented in the study population raises concerns about the utility and generalizability of GRADE study findings and its impact on diabetes management, underscoring the important complementary insights that can be gleaned from analyses of real-world data (which can be designed to use more pragmatic and generalizable eligibility criteria) as adjuncts to RCTs.

We met our objective to conduct all analyses prior to publication of GRADE trial findings, and it will be important to ultimately compare our findings to those in GRADE. The greater effectiveness of liraglutide compared to both glimepiride and sitagliptin is consistent with prior studies.^30 32-34^ Additionally, subgroup analyses demonstrating greater effectiveness of liraglutide among patients with elevated baseline HbA_1c_ and in younger patients, generated important hypotheses regarding the optimal use of liraglutide (and potentially other GLP-1RA) in clinical practice to be explored in future research. When the GRADE trial was conceived, drugs’ ability to lower HbA_1c_ was at the forefront of clinical decision making when choosing glucose-lowering therapy. Similarly, the SGLT2i class of glucose-lowering medications had not yet been incorporated into practice and therefore was excluded as a comparator therapy when the GRADE trial was conceived and designed. In addition, contemporary clinical practice guidelines increasingly focus on the impacts of glucose-lowering therapies on hard outcomes that are important to patients such macrovascular and microvascular complications and death, beyond HbA_1c_.^35^ Indeed, most recent clinical practice guidelines recommend consideration of GLP-1RA and SGLT2i even as first-line therapies and independent of the HbA_1c_ level among patients at high risk for ASCVD, kidney disease, and heart failure.^14^ For these outcomes, there is robust evidence favoring liraglutide (of the drug classes examined) in patients at high risk for ASCVD,^36 37^ further underscoring its advantage. It will be important, in future research, to compare the effectiveness of glycemic control achieved by GLP-1RA with that of SGLT2i, as SGLT2i are similarly recommended for patients at high risk for cardiovascular disease, kidney disease, and heart failure.^14^

Our study is strengthened by application of advanced analytic methods that account for measured differences between treatment arms that otherwise confound analyses and preclude causal inference. The GBM-based models for the PS are more flexible and less sensitive to model misspecification compared to logistic regression. The large and diverse patient population within OLDW made emulation efforts uniquely possible despite the narrow eligibility criteria specified by GRADE.

### Limitations

Despite rigorous causal inference analytic methods, observational studies are inevitably subject to residual confounding. For the metabolic endpoints, there was evidence of non-proportional hazards, which makes the single summary hazard ratio calculated from the Cox proportional hazards an imperfect estimate for the time-varying risk. However, with the goal of emulating the GRADE trial, where the statistical analysis plan was to estimate single summary hazard ratios, we report the same estimate in the emulation. Nevertheless, we were unable to operationalize every component of GRADE’s eligibility criteria and end points. For example, we did not require confirmatory HbA_1c_ results to meet the metabolic endpoints and were not able to maintain the same standard timeframe for HbA_1c_ ascertainment as specified in GRADE. Additionally, while the GRADE trial analyses were conducted using the ITT principle, we a priori chose to use per-protocol analysis for the metabolic endpoints because in the absence of randomization, reasons for changing a treatment typically depend on post-initiation factors that could confound the association between the treatment group and the outcome. While advanced statistical methods can account for post-baseline differences between groups in key characteristics, these methods require accurate estimation of the reasons to stop or change treatment, and such estimation is not feasible in this setting using claims data. Duration of follow-up was also different among the treatment arms, which is unavoidable when studying real-world practice patterns. In particular, a higher proportion of patients initiating liraglutide filled only one cycle of treatment before either switching to a different treatment or not refilling their prescription, potentially due to poor tolerability, subcutaneous administration, or high cost.

Not all patients with claims data in OLDW have available laboratory data, as results are available for a subset of patients based on data sharing agreements between OptumLabs and commercial laboratories. However, laboratory results availability is independent of treatment regimen, and we do not expect it to bias our analyses. The schedule of HbA_1c_ testing in real-world practice is contingent on the patient’s current HbA_1c_ level, the clinician’s anticipation of changing HbA_1c_ levels, and the patient’s ability to access care. This may have confounded study results by delaying the time to HbA_1c_ reassessment and reaching the study endpoint in patients with low baseline HbA_1c_ or with barriers to care. Our evaluation could not account for inclusion and exclusion criteria that could not be operationalized using claims data, including medications obtained without insurance coverage (e.g., obtained through a low-cost generic program,^38^ patient assistance program, or a sample), comorbidities that were not coded and billed in a clinical encounter, and family history information. However, prior studies found the likely number of glucose-lowering medications missing from claims to be low.^39^ Finally, the study cohort was comprised of patients with private and Medicare Advantage health plans, such that results may not fully generalize to patients with public health plans or those without insurance coverage.

## CONCLUSIONS

Better understanding of the comparative effectiveness and safety of second-line glucose-lowering medications is urgently needed to inform shared decision-making in diabetes. Analytic methods such as those implemented in this study, and in the parallel emulation of PRONOUNCE,^18^ can be leveraged for more timely evaluations of drug effectiveness and safety as long as the treatments being considered are already being used in clinical practice. Indeed, work is currently under way to examine the comparative effectiveness of sulfonylurea, GLP-1RA, DPP-4i, and SGLT2i drugs with respect to ASCVD and other hard outcomes among patients at moderate risk for ASCVD using observational data from real-world practice.^40^ Ultimately, the population included in this study and our findings should be compared to those of the GRADE trial, once published in peer-reviewed literature, to assess the fidelity and generalizability of results and to deepen our understanding of the use of real-world data to emulate clinical trials.

## Data Availability

OLDW data are available for research through a virtual data warehouse. The authors are not able to distribute the data. Study protocol, code sets, and statistical analysis plan are available online.

## Data Sharing

This study was conducted using de-identified claims data from OptumLabs Data Warehouse. Raw data are not publicly available. The study protocol is available online.^23^

