## Supplemental Files for "Emulating the GRADE Trial Using Real-World Data"

**Online Only Supplemental Material**

Figure S5. Cumulative risks of secondary metabolic failure in propensity score weighted patients…………………………………………………………………………………………...24

### Supplemental Methods.

#### ***Source data for individual-level socioeconomic information***

Socioeconomic status data available in OptumLabs Data Warehouse (OLDW) is sourced from a national supplier of consumer marketing data, which includes consumer-specific demographic, behavioral, and lifestyle information. Much like administrative claims, the data have been collected for purposes other than research and variable imputation is conducted by the Data Supplier as appropriate. Socioeconomic variables included in our analyses are race, ethnicity, and annual household income. Race and ethnicity are imputed based on a model run by the Data Supplier using an individual’s name (first, last, middle) and geographic location, then categorized into five race values in OLDW: W (White), B (Black), H (Hispanic), A (Asian), and U (Unknown). The Data Supplier imputes estimated household income based on a model using both public and private consumer data. This variable is assigned at the household level where a “household” is defined as individuals with the same surname living at the same street address.

#### Propensity score estimation with multiple treatments using generalized boosted models

Let $T_{i}\in\{1, 2, \ldots, M\}$ indicate the treatment group for patient *i* out of the *M* treatment groups, and $X_{i}$ be the collection of baseline covariates. Then the propensity score can be defined as $\mathrm{ps}_{t}= Pr \left( T=t \right|X)$. For each treatment group, a logistic regression model was estimated for the given treatment versus all others. Regular logistic regression could be used to estimate the individual propensity scores, but requires the functional form of the baseline covariates in the regression model to be close to the truth. An alternative is to estimate the functional form of the baseline covariates using a boosting model as described in McCaffrey et al (2013).^1^ The collection of generalized boosted models was estimated using the *gbm* package in R.^2^ The base model in the boosted ensemble was a regression tree with interaction depth 3. The optimal number of trees in the boosted ensemble was selected based on the number of trees that minimized the maximum standardized effect size between treatment groups across the baseline covariates.

### Figure S1. Cohort creation prior to application of GRADE Trial eligibility criteria.

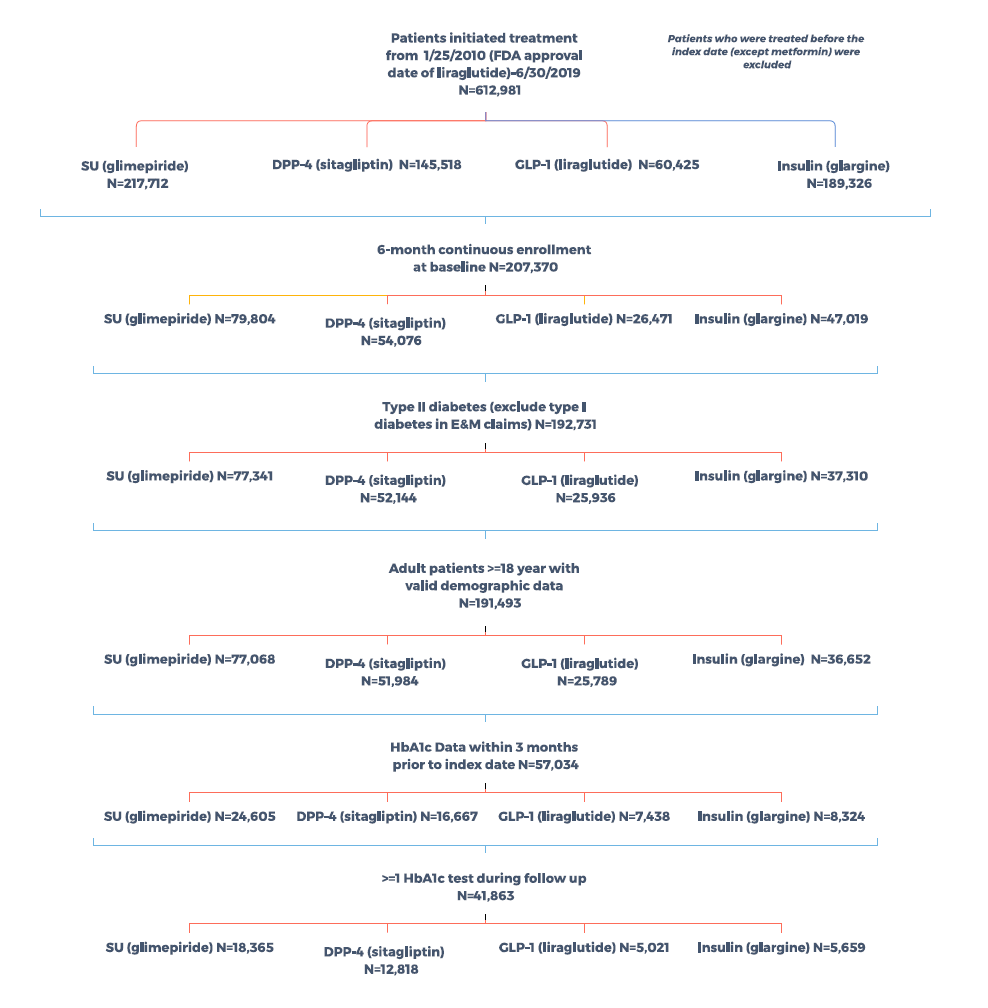

### Table S1. Eligibility criteria for the GRADE trial and its OLDW emulation.

| **GRADE Eligibility Criteria** | **Operational Definition in OLDW** |
| --- | --- |
| **Inclusion criteria** |  |
| Men or women ≥30 years of age at time of diabetes diagnosis.  For American Indians: ≥20 years of age at time of diagnosis | Age ≥30 years at index date (*American Indians are classified as “other” race/ethnicity in OLDW*) |
| Duration of diagnosed diabetes: <10 years determined as accurately as possible on the basis of available records at screening | N/A (*We are not able to determine the duration of diagnosed diabetes*) |
| HbA_1c_ criteria (at final run-in visit, ~2 weeks before randomization): 6.8–8.5% (51–69 mmol/ mol) | HbA_1c_ result in the range (6.8–8.5%) within 3 months (inclusive) of the index date |
| Taking a daily dose of ≥1,000 mg metformin for a minimum of 8 weeks at final run-in | Adherent to metformin (no dose requirement) for ≥ 8 weeks before the index date |
| Willingness to administer daily subcutaneous injections, take a second diabetes drug after randomization, potentially initiate insulin, intensify insulin therapy if study metabolic goals are not met, and perform self-monitoring of blood glucose | N/A |
| A negative pregnancy test for all women of childbearing potential (i.e., premenopausal, not surgically sterile) | No diagnosis or procedure codes at any position for pregnancy within 6 months before and inclusive of the index date |
| Provision of signed and dated informed consent before any study procedures | N/A |
| **Exclusion criteria** |  |
| Suspected type 1 diabetes (lean with polyuria, polydipsia, and weight loss with little response to metformin) or secondary diabetes resulting from specific causes (e.g., previously diagnosed monogenic syndromes, pancreatic surgery, pancreatitis) | Diagnosis codes for type 1 diabetes at any position of any E&M encounter at any time during enrollment period prior index date  *(Excluded as part of inclusion criteria)* |
| Current or previous (within past 6 months) treatment with any diabetes drug or glucose lowering medication other than metformin, including short-term insulin use during hospitalization | Fill of any diabetes medications (except metformin) within 6 months before index date  *(Excluded as part of inclusion criteria)* |
| More than 10 years of treatment with metformin at time of randomization | N/A |
| History of intolerance, allergy, or other contraindications to any of the proposed study medications | N/A |
| A life-threatening event within 30 days before screening or currently planned major surgery | N/A |
| Any major cardiovascular event in previous year, including history of myocardial infarction, stroke, or vascular procedure, such as coronary artery or peripheral bypass grafting, stent placement (peripheral or coronary), or angioplasty | Diagnosis and procedure codes at any position for major cardiovascular events within one year before and including the index date |
| Plans for pregnancy during the course of the study for women of childbearing potential | Diagnosis and procedure codes at any position for pregnancy within 6 months before and inclusive of the index date  *(Excluded as part of inclusion criteria)* |
| History of or planning for bariatric surgery, including banding procedures or surgical gastric and/or intestinal bypass | Diagnosis and procedure codes at any position for bariatric surgery within 6 months before and including the index date |
| History of congestive heart failure (New York Heart Association class III or IV) | Diagnosis codes for congestive heart failure at any position within 6 months before and inclusive of the index date |
| History of conditions that are specific contraindications to any of the study medications | N/A |
| Serum creatinine level ≥1.4 mg/dL in women and ≥1.5 mg/dL in men or end-stage renal disease requiring renal replacement therapy | Serum creatinine level ≥1.4 mg/dL in women and ≥1.5 mg/dL in men; or kidney transplant codes at baseline; or ≥3 dialysis codes at baseline; or Diagnosis code for ESRD within 6 months before and inclusive of the index date, all codes were pulled at any position |
| History of cancer, other than nonmelanoma skin cancer, that required therapy in the 5 years before randomization | Diagnosis codes at any position for cancer (except nonmelanoma skin cancer) within 6 months before and inclusive of the index date |
| Treatment with oral or systemic glucocorticoids (other than short-term treatment, e.g., for poison ivy) or disease likely to require periodic or regular glucocorticoid therapy (inhaled steroids allowed) | Adherent to oral glucocorticoids for ≥3 months at baseline |
| Treatment with atypical antipsychotics | A prescription of atypical antipsychotics within 3 months before or on the index date |
| Clinically or medically unstable with expected survival <1 year | N/A |

Table S2. Code sets used to ascertain comorbidities included in eligibility criteria, baseline covariates, or outcomes. All baseline covariates were assessed at all positions in diagnosis and procedure codes.

| **Comorbidity** | **ICD-9 Codes** | **ICD-10 codes** | **CPT codes** | **Revenue codes** |
| --- | --- | --- | --- | --- |
| **Bariatric surgery** | V45.86, 43.82, 43.89, 44.38, 44.39, 44.68, 44.95, 44.69 | Z98.84, 0DQ64ZZ, 0DQ60ZZ, 0DB80ZZ, 0D16078, 0D16479, 0DV64CZ, 0D190Z9, 0DB60ZZ | 43644, 43645, 43770, 43775, 43842, 43843, 43845, 43846, 43847, S2082, S2085 |  |
| **Cancer (except non-melanoma skin cancer)** | 14x.xx, 15x.xx, 160.x-165.x, 170.x-172.x, 174.x-176.x, 179-189.x, 190.x-195.x, 199.xx-208.xx (except 203.x1, 204.x1, 205.x1, 206.x1, 207.x1, 208.x1) | C00.x-C14.x, C15.x-C26.x, C3x.xx, C40.xx-C41.x, C43.x, C4A.xx, C45.x-C49.xx, C50.xxx, C51.x-C58, C60.x-C63.x, C64.x-C68.x, C69.xx-C72.x, C73-C76.x, C7A.xx, C80.xx-C96.x (except C90.x1, C91.x1, C92.x1, C93.x1, C94.x1, C95.x1) |  |  |
| **Coronary artery disease** | 429.2, 410.x, 411.x, 412.x, 413.x, 414.x | I20.x, I21.x, I22.x, I23.x, I24x, I25.x |  |  |
| **Coronary artery bypass grafting surgery, percutaneous coronary intervention, lower extremity revascularization** | 36.1x, V45.81, 414.02, 414.03, 414.04, 414.05  V45.82, 36.0x, 00.66  39.25, 39.29, 38.08, 38.16, 38.18, 38.38, 38.48, 38.68, 38.88, 3950, 39.90, 00.55, 84.3, 84.1x | 02100x, 02110x, 02120x, 02130x, Z95.1, T82.21x, I25.7x, I25.810, I25.812  Z98.61, Z95.5, 0270x, 0271x, 0272x, 0273x, 02C0x, 02C1x, 02C2x, 02C3x  041x, 047x, 04Bx, 04Cx, 04Lx, 04Px, 04Rx (4^th^ letter C-Y, except G, I, O, X) | 33510, 33511, 33512, 33513, 33514, 33516, 33517, 33518, 33519, 33521, 33522, 33523, 33533, 33534, 33535, 33536, 4110F  92920, 92921, 92924, 92925, 92928, 92929, 92933, 92934, 92937, 92938, 92941, 92943, 92944, 92980, 92981, 92982, 92984, 92995, 92996, 92975, 92977 |  |
| **Cerebrovascular disease** | 430, 431, 432.x, 433.xx, 434.xx 435.x, 436, 437.x, 438.xx, V12.54 | G45.0, G45.1, G45.2, G45.8, G45.9, G46.x, I60.xx, I61.x (except I61.0), I62.xx, I63.xxx, I65.xx, I66.xx, I67.8x (except I67.83, I67.84), I67.9, I69.xxx, Z86.73 |  |  |
| **Chronic kidney disease** | 403.01, 403.11, 403.91, 404.02, 404.03, 404.12, 404.13, 404.92, 404.93, 585.3, 585.4, 585.5, 585.6, 792.5, 996.81, V42.0, V45.1, V45.11, V45.12, V56, V56.0, V56.1, V56.2, V56.3, V56.31, V56.32, V56.8 | I12.0, I13.11, I13.2, I95.3, N18.3, N18.4, N18.5, N18.6, R88.0, T86.1, T86.10, T86.11, T86.12, T86.13, T86.19, Y84.1, Z48.22, Z49, Z49.0, Z49.01, Z49.02, Z49.3, Z49.31, Z49.32, Z91.15, Z94.0, Z99.2, T81502x, T81512x, T81522x, T81532x, T81592x, T85611x, T85621x, T85631x, T85651x, T8571x |  |  |
| **End-stage kidney disease, dialysis, transplantation** | 403.01, 403.11, 403.91, 404.02, 404.03, 404.12, 404.13, 404.92, 404.93,  585.5, 585.6, 792.5,  996.81, V42.0, V45.1,  V45.11, V45.12, V56.x, V56.0, V56.1, V56.2, V56.3, V56.31, V56.32, V56.8  39.95, 54.98, 55.53, 55.6, 55.69 | I12.0, I13.11, I13.2, I95.3, N185, N186, R88.0, T81.502x, T81.512x, T81.522x, T81.532x, T81.592x, T85.611x, T85.621x, T85.631x, T85.651x, T85.71x, T86.1, T86.10, T86.11, T86.12, T86.13, T86.19, Y84.1, Z48.22, Z49, Z49.0, Z49.01, Z49.02, Z49.3, Z49.31, Z49.32, Z91.15, Z94.0, Z99.2  0TT00ZZ, 0TT04ZZ, 0TT10ZZ, 0TT14ZZ, 0TT30ZZ, 0TT34ZZ, 0TT37ZZ, 0TT38ZZ, 0TT40ZZ, 0TT44ZZ, 0TY00Z0, 0TY00Z1, 0TY00Z2, 0TY10Z0, 0TY10Z1, 0TY10Z2, 3E1M39Z, 5A1D00Z, 5A1D60Z | 50340, 50360, 50365, 50370, 90935, 90937, 90940, 90945, 90947, 90957, 90958, 90959, 90960, 90961, 90962, 90965, 90966, 90969, 90970, 90999, G0257, S9335, 99512 | 0800, 0801, 0802, 0803, 0804, 0805, 0806, 0807, 0808, 0809, 0820, 0821, 0822, 0823, 0824, 0825, 0826, 0827, 0828, 0829, 0830,  0831, 0832, 0833, 0834, 0835, 0836, 0837, 0838, 0839, 0840, 0841, 0842, 0843, 0844, 0845, 0846, 0847, 0848, 0849, 0850, 0851, 0852, 0853, 0854, 0855, 0856, 0857, 0858, 0859, 0880, 0881, 0882, 0883, 0884, 0885, 0886, 0887, 0888, 0889 |
| **Hyperglycemia** | 250.10, 250.11, 250.12, 250.13, 250.20, 250.21, 250.22, 250.23 | E10.10, E10.11, E11.10, E11.11, E13.10, E13.11, E11.00, E11.01, E13.00, E13.01 |  |  |
| **Heart failure** | 398.91, 402.01, 402.11, 402.91, 404.01, 404.03, 404.11, 404.13, 404.91, 404.93, 428.xx | I09.81, I11.0, I13.0, I13.2, I50.xx |  |  |
| **Hypoglycemia** | 251.0, 251.1, 251.2, 962.3, 250.8x (for 250.8x: if no concurrent 259.8, 272.7, 681.xx, 682.xx, 686.9x, 707.1x-707.2x, 707.8, 707.9, 709.3, 730.0x-730.2x, 731.8) | E10.641, E10.649, E11.641, E11.649, E13.641, E13.649, E16.0, E16.1, E16.2, T38.3X1A, T38.3X1D, T38.3X1S, T38.3X2A, T38.3X2D, T38.3X2S, T38.3X3A, T38.3X3D, T38.3X3S, T38.3X4A, T383X4D, T38.3X4S, T38.3X5A, T383X5D, T38.3X5S |  |  |
| **Myocardial infarction** | 410.x, 412.x | I21.x, I22.x, I23.x, I252.x |  |  |
| **Nephropathy** | 593.9, 586, 250.4x, 249.4x, 580.x, 581.x, 582.x, 583.x, 585.x | E08.21, E08.22, E08.29, E09.21, E09.22, E09.29, E10.21, E10.22, E10.29, E11.21, E11.22, E11.29, E13.21, E13.22, E13.29, N19, N00.x, N03.x, N04.x, N05.x, N18.x |  |  |
| **Neuropathy** | 357.2, 337.1, 356.9, 358.1, 458.0, 536.3, 564.5, 596.54, 713.5, 951.0, 951.1, 951.3, 250.6x, 249.6x, 337.0x, 354.x, 355.x | G90.09, G90.8, G90.9, G99.0, G60.9, G73.3, G90.01, I95.1, K31.84, K59.1, N31.9, E08.4x, E09.4x, E10.4x, E11.4x, E13.4x, G56.x, G57.x, H49.x, M14.6x, S04.x |  |  |
| **Peripheral vascular disease** | 442.3, 440.21, 443.81, 443.9, 892.1, 040.0, 444.22, 785.4, 250.7x, 249.7, 707.1x | E08.51, E09.51, E10.51, E11.51, E13.51, E08.59, E09.59, E10.59, E11.59, E13.59, E08.621, E09.621, E10.621, E11.621, E13.621, I72.4, I73.89, I73.9, A48.0, I74.3, I96, E08.52, E09.52, E10.52, E11.52, E13.52, I70.21x, S91.3x, L97.x |  |  |
| **Pneumonia** | 487.0, 480.x, 481.x 482.x, 483.x, 485.x, 486.x | A01.03, A02.22, A37.01, A37.11, A37.81, A37.91, A50.04, A54.84, B01.2, B05.2, B06.81, B77.81, J09.X1, J13, J14, J17, J84.2, J85.1, J85.2, J95.851, J10.0x, J11.0x, J12.x, J15.x, J16.x, J18.x, J84.11x |  |  |
| **Retinopathy** | 362.01, 362.03, 362.04, 362.05, 362.06, 362.07, 362.53, 362.81, 362.82, 362.83, 362.02, 379.23, 250.5x, 249.5x, 362.1x, 361.x, 369.x | H35.9, E08.3x, E09.3x, E10.3x, E11.3x, E13.3x, H35.0x, H35.35x, H35.6x, H35.8x, H33.x, H54.x, H43.1x |  |  |
| **Stroke** | 433.01, 433.11, 433.21, 433.31, 433.81, 433.91, 434.01, 434.11, 434.91, 436, 430, 431 | I63.x, I60.x, I61.x, I69.0x, I69.1x, I69.3x |  |  |
| **Type 1 diabetes** | 250.x1, 250.x3 | E10.x |  |  |

### Table S3. Medications included in eligibility criteria or baseline covariates.

| **Medication Class** | **Included Medications** |
| --- | --- |
| **Anti-psychotics** | Aripiprazole, Asenapine, Asenapine Maleate, Brexpiprazole, Cariprazine HCL, Clozapine, Lurasidone HCL, Olanzapine, Olanzapine/Fluoxetine HCL, Paliperidone, Quetiapine Fumarate, Risperidone, Ziprasidone HCL |
| **ACE inhibitors** | Benazepril HCL, Captopril, Enalapril Maleate, Fosinopril Sodium, Lisinopril, Perindopril Erbumine, Quinapril HCL, Ramipril, Trandolapril, Moexipril HCL, Benazepril/Hydrochlorothiazide, Captopril/Hydrochlorothiazide, Enalapril/Hydrochlorothiazide, Fosinopril/Hydrochlorothiazide, Lisinopril/Hydrochlorothiazide, Moexipril/Hydrochlorothiazide, Quinapril/Hydrochlorothiazide, Amlodipine Besylate/Benazepril, Enalapril Maleate/Diltiazem, Enalapril Maleate/Felodipine, Perindopril Arginine/Amlodipine Bes, Trandolapril/Verapamil HCL |
| **Angiotensin receptor blockers** | Candesartan Cilexetil, Irbesartan, Losartan Potassium, Olmesartan Medoxomil, Telmisartan, Valsartan, Azilsartan Medoxomil, Eprosartan Mesylate, Azilsartan Med/Chlorthalidone, Olmesartan/Hydrochlorothiazide, Candesartan Cilexetil/HCTZ, Candesartan/Hydrochlorothiazide, Eprosartan Mesylate/HCTZ, Eprosartan/Hydrochlorothiazide, Irbesartan/Hydrochlorothiazide, Losartan Potassium/HCTZ, Losartan/Hydrochlorothiazide, Olmesartan Medoxomil/HCTZ, Olmesartan/Hydrochlorothiazide, Telmisartan/HCTZ, Telmisartan/Hydrochlorothiazide, Valsartan/Hydrochlorothiazide |
| **Warfarin and direct oral anti-coagulants (DOAC)** | Warfarin, Dabigatran, Rivaroxaban, Apixaban, Edoxaban |
| **Glucocorticoids** | Hydrocortisone, Cortisone Acetate, Prednisone, Prednisolone, Methylprednisolone, Triamcinolone, Dexamethasone, Betamethasone |
| **Lipid-lowering medications** (not statins) | Alirocumab, Evolocumab, Ezetimibe, Mipomersen, Lomitapide, Bezafibrate, Clofibrate, Fenofibrate, Fenofibric Acid, Gemfibrozil, Cholestyramine, Colesevelam, Colestipol |
| **Peripheral neuropathy medications** | Gabapentin, Pregabalin, Duloxetine |
| **Statins** | Atorvastatin, Rosuvastatin, Simvastatin, Pravastatin, Lovastatin, Fluvastatin, Pitavastatin |
| **Biguanide** | Metformin |
| **Sulfonylurea** | Glimepiride, Glipizide, Glyburide |
| **DPP-4 inhibitor** | Alogliptin, linagliptin, sitagliptin, saxagliptin |
| **GLP-1 receptor agonist** | Exenatide, liraglutide, albiglutide, dulaglutide, semaglutide, lixisenatide |
| **SGLT2 inhibitor** | Canagliflozin, empagliflozin, dapagliflozin, ertugliflozin |
| **Thiazolidinedione** | Pioglitazone, Rosiglitazone |
| **Basal insulin** | NPH, Isophane, Zinc, Glargine, Detemir, Degludec |
| **Bolus insulin** | Regular insulin, Beef insulin, Pork insulin, Aspart, Lispro, Glulisine |
| **Other glucose-lowering medications** | Nateglinide, Repaglinide, Pramlintide, Acarbose, Miglitol |

Table S4. Frequency and timing of HbA_1c_ tests in each study arm. Because hemoglobin A_1c_ (HbA_1c_) testing frequency and intervals vary in real-world practice, and differential availability of test results between study treatment arms may bias the results, we examined the frequency, intervals, and total number of available HbA_1c_ tests within each treatment arm. Importantly, clinical practice guidelines advise that testing frequency is determined by the patient’s glycemic control (i.e., HbA_1c_ level and glycemic variability) rather than their treatment regimen.^3^

| **Medication** | **N** | **Mean** | **Median** | **Lower Quartile** | **Upper Quartile** | **Min** | **Max** |
| --- | --- | --- | --- | --- | --- | --- | --- |
| **Days between HbA_1c_ testing** | | | | | | | |
| Glimepiride | 4318 | 156 | 123 | 92 | 186 | 1 | 2196 |
| Liraglutide | 690 | 155 | 119 | 90 | 184 | 1 | 1619 |
| Sitagliptin | 2993 | 158 | 121 | 92 | 188 | 1 | 2098 |
| **Days between index date and first HbA_1c_ after the index date** | | | | | | | |
| Glimepiride | 4318 | 161 | 116 | 86 | 184 | 1 | 2196 |
| Liraglutide | 690 | 149 | 96 | 72 | 174 | 1 | 1497 |
| Sitagliptin | 2993 | 157 | 106 | 81 | 182 | 1 | 2098 |
| **Number of HbA_1c_ tests after the index date, before censoring per protocol** | | | | | | | |
| Glimepiride | 3017 | 2 | 2 | 1 | 4 | 1 | 27 |
| Liraglutide | 387 | 2 | 1 | 1 | 3 | 1 | 23 |
| Sitagliptin | 1898 | 2 | 2 | 1 | 3 | 1 | 20 |
| **Number of HbA_1c_ tests after the index date, before censoring per intention to treat** | | | | | | | |
| Glimepiride | 4318 | 4 | 3 | 2 | 6 | 1 | 33 |
| Liraglutide | 690 | 3 | 3 | 2 | 6 | 1 | 29 |
| Sitagliptin | 2993 | 4 | 3 | 2 | 6 | 1 | 33 |

Table S5. HbA_1c_ test intervals in each study arm, stratified by baseline HbA_1c_ level. Because hemoglobin A_1c_ (HbA_1c_) testing frequency is informed by the patient’s baseline HbA_1c_ level,^3^ we examined the number of days in between sequential HbA_1c_ results in each treatment arm stratified by their baseline HbA_1c_.

| **Medication** | **Baseline HbA_1c_** | **N** | **Mean** | **Median** | **Lower Quartile** | **Upper Quartile** |
| --- | --- | --- | --- | --- | --- | --- |
| Glimepiride | <7.0% | 295 | 150 | 121 | 91 | 185 |
|  | ≥7.0% | 4023 | 156 | 123 | 92 | 187 |
| Liraglutide | <7.0% | 100 | 174 | 135 | 91 | 217 |
|  | ≥7.0% | 590 | 152 | 118 | 90 | 182 |
| Sitagliptin | <7.0% | 289 | 163 | 120 | 91 | 190 |
|  | ≥7.0% | 2704 | 158 | 121 | 92 | 188 |

### Table S6. Secondary outcomes included in the analyses.

1) ***Metabolic endpoints***
   (1) Proportion of subjects among those randomized to each treatment that has reached the primary metabolic outcome over time
   (2) Time to secondary metabolic failure (HbA_1c_ >7.5%) after having reached the primary outcome, while receiving the assigned regimen.
   (3) Proportion of subjects among those randomized to each treatment that has reached secondary metabolic failure over time.
   (4) Time to the need for insulin therapy while being treated with the assigned regimen.
   (5) Proportion of subjects among those randomized to each treatment group that has initiated insulin therapy over time.
   (6) Number of emergency department visits and/or hospitalizations for hypoglycemia per 1000 patients per year, ascertained using primary diagnosis codes in ED/hospitalization claims
2) ***Microvascular outcomes***
(1) Incidence* of end-stage kidney disease, ascertained using diagnosis and procedure codes at any position in E&M claims (note: this outcome was not present in GRADE, which examined progression of microalbuminuria or worsening of eGFR)
(2) Incidence of diabetic retinopathy or documentation of retinal photocoagulation for diabetic retinopathy, ascertained using diagnosis and procedure codes at any position in E&M claims.
(3) Incidence of peripheral neuropathy, ascertained using diagnosis codes at any position in E&M claims.
3) ***Cardiovascular outcomes***
(1) Incidence of major adverse cardiovascular events (MACE: death, nonfatal myocardial infarction, nonfatal stroke), MI and stroke were ascertained using primary and first secondary diagnosis codes in hospitalization claims. Death was ascertained from OLDW, where it is derived from the Social Security Administration Death Master File, deceased status from electronic medical records, death as a reason for disenrollment, death indicated by inpatient discharge status, obituary information, and Medicare Advantage beneficiary report information.
(2) Incidence of other cardiovascular events including unstable angina requiring hospitalization or revascularization, ascertained using primary and first secondary diagnosis and procedure codes in hospitalization claims
(3) Incidence of congestive heart failure requiring hospitalization, ascertained using primary diagnosis codes in hospitalization claims.
5) ***Safety outcomes (adverse events)***
(1) Incidence of pancreatitis, ascertained using diagnosis codes at any position in ED/hospitalization claims
(2) Incidence of pancreatic and medullary thyroid cancer, ascertained using diagnosis codes at any position in E&M claims
(3) Incidence of cancer (except non-melanoma skin cancer) , ascertained using diagnosis codes at any position in E&M claims
6) ***Other outcomes***
   (1) All-cause mortality
   (2) Any hospital admission, ascertained using revenue codes

*Incidence: no codes at any position for this category in the past 12 months

### Figure S2. Distribution of stabilized propensity score weights, stratified by drug.

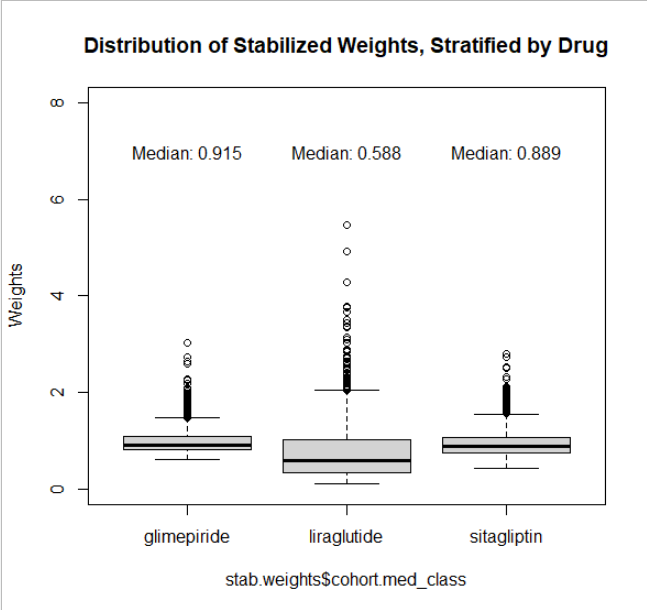

Figure S3. Censoring approaches. For the primary outcome (primary metabolic failure) and the secondary outcomes of secondary metabolic failure and initiation of insulin, patients were censored when the assigned treatment drug was discontinued (point 3) or at death, disenrollment, or end of the study period (point 4), whichever came first. For all other secondary outcomes, we censored at the time of death, disenrollment, or end of the study period (point 4). As a sensitivity analysis (“as treated approach”), patients were censored when they added a different medication class (point 2), discontinued the assigned drug (point 3), or at death, disenrollment, or end of the
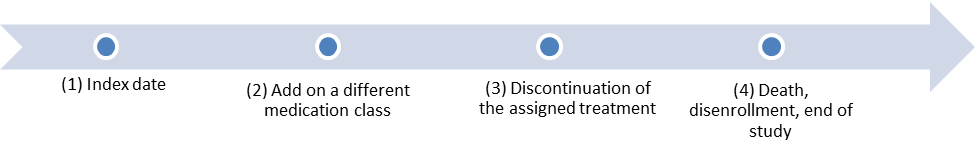
study period (point 4), whichever came first.

Rationale: For the primary analysis, a patient was right censored if they discontinued their initial study treatment (“per protocol” approach), but we did not consider adding an additional antidiabetic treatment to the current initiated treatment as sufficient to censor the individual from follow-up. Since the primary outcome defined in the GRADE protocol was “time to an HbA_1c_ value ≥7% while subjects are treated with the maximally tolerated doses of both metformin and the randomly assigned medication,” we similarly continued follow patients as long as they were still taking their initiated study treatment. However, because adding an additional treatment may indicate a different clinical course, a sensitivity analysis was performed to censor at these time points to evaluate sensitivity of the treatment group comparisons (“as treated” approach).

Table S7. Patients excluded from analyses due to not meeting GRADE Trial eligibility criteria. GRADE eligibility criteria were applied to the base population of adults (age ≥18 years) with type 2 diabetes included in OptumLabs Data Warehouse (OLDW) who had available hemoglobin A_1c_ data and the requisite baseline enrollment period.

| **GRADE Eligibility Criteria using the Operational Definition in OLDW** | **Overall**  **(N =41,863)** | **Glargine**  **(N =5659)** | **Glimepiride**  **(N =18365)** | **Liraglutide**  **(N =5021)** | **Sitagliptin**  **(N =12,818)** |
| --- | --- | --- | --- | --- | --- |
| Total % of patients eligible for GRADE | 8252 (19.7%) | 251 (4.4%) | 4318 (23.5%) | 690 (13.7%) | 2993 (23.3%) |
| Total % of patients ineligible for GRADE | 33611 (80.3%) | 5408 (95.6%) | 14047 (76.5%) | 4331 (86.3%) | 9825 (76.7%) |
| **Patients who did NOT meet the inclusion criterion below (%)** | | | | | |
| Age ≥30 years at index date | 473 (1.1%) | 143 (2.5%) | 124 (0.7%) | 135 (2.7%) | 71 (0.6%) |
| Have HbA_1c_ between 6.8–8.5% within 3 months of index date | 24529 (58.6%) | 4591 (81.1%) | 9701 (52.8%) | 3606 (71.8%) | 6631 (51.7%) |
| Adherent to metformin for ≥8 weeks before the index date | 20376 (48.7%) | 3874 (68.5%) | 7997 (43.5%) | 2749 (54.8%) | 5756 (44.9%) |
| No diagnosis or procedure codes for pregnancy at baseline | 98 (0.2%) | 56 (1.0%) | 12 (0.1%) | 19 (0.4%) | 11 (0.1%) |
| **Patients who met one of the following exclusion criteria (%)** | | | |  |  |
| Diagnosis and procedure codes for major cardiovascular events within one year before index date | 3509 (8.4%) | 850 (15.0%) | 1467 (8.0%) | 175 (3.5%) | 1017 (7.9%) |
| Diagnosis and procedure codes for bariatric surgery at baseline | 139 (0.3%) | 26 (0.5%) | 36 (0.2%) | 51 (1.0%) | 26 (0.2%) |
| Diagnosis codes for congestive heart failure at baseline | 2378 (5.7%) | 609 (10.8%) | 967 (5.3%) | 142 (2.8%) | 660 (5.1%) |
| Serum creatinine level >1.4 mg/dL in women and >1.5 mg/dL in men; or kidney transplant codes at baseline; or ≥3 dialysis codes at baseline; or diagnosis code for ESRD at baseline | 2073 (5.0%) | 429 (7.6%) | 919 (5.0%) | 98 (2.0%) | 627 (4.9%) |
| Diagnosis codes for cancer (except nonmelanoma skin cancer) at baseline | 2570 (6.1%) | 464 (8.2%) | 1187 (6.5%) | 165 (3.3%) | 754 (5.9%) |
| Adherent to oral glucocorticoids for ≥3 months at baseline | 599 (1.4%) | 145 (2.6%) | 252 (1.4%) | 42 (0.8%) | 160 (1.2%) |
| A prescription of atypical antipsychotics within 3 months before or on the index date | 897 (2.1%) | 173 (3.1%) | 337 (1.8%) | 142 (2.8%) | 245 (1.9%) |

### Table S**8**. **Complete list of medications included in the final cohort**.

|  | **Frequency** | **Percent** |
| --- | --- | --- |
| Glimepiride | 4314 | 53.92 |
| Liraglutide | 690 | 8.62 |
| Pioglitazone HCL/Glimepiride | 3 | 0.04 |
| Pioglitazone/Glimepiride | 1 | 0.01 |
| Sitagliptin Phos/Metformin HCL | 849 | 10.61 |
| Sitagliptin Phosphate | 2144 | 26.80 |

### Table S9. Baseline characteristics of patients in the four treatment arms prior to weighting.

|  | **Glargine (N=251)** | **Glimepiride (N=4318)** | **Liraglutide (N=690)** | **Sitagliptin (N=2993)** | **Total (N=8252)** | **Largest SMD** |
| --- | --- | --- | --- | --- | --- | --- |
| **Age, mean (SD)** | 60.2 (12.4) | 62.9 (11.0) | 54.9 (9.8) | 61.9 (11.2) | 61.8 (11.2) | 0.67 |
| **Age group, years** |  |  |  |  |  | 0.81 |
| 30-44 | 30 (12.0%) | 268 (6.2%) | 97 (14.1%) | 237 (7.9%) | 632 (7.7%) |  |
| 45-54 | 49 (19.5%) | 726 (16.8%) | 250 (36.2%) | 538 (18.0%) | 1563 (18.9%) |  |
| 55-64 | 65 (25.9%) | 1125 (26.1%) | 221 (32.0%) | 783 (26.2%) | 2194 (26.6%) |  |
| 65-74 | 81 (32.3%) | 1621 (37.5%) | 110 (15.9%) | 1095 (36.6%) | 2907 (35.2%) |  |
| ≥75 | 26 (10.4%) | 578 (13.4%) | 12 (1.7%) | 340 (11.4%) | 956 (11.6%) |  |
| **Gender** |  |  |  |  |  | 0.19 |
| Female | 132 (52.6%) | 1982 (45.9%) | 410 (59.4%) | 1499 (50.1%) | 4023 (48.8%) |  |
| Male | 119 (47.4%) | 2336 (54.1%) | 280 (40.6%) | 1494 (49.9%) | 4229 (51.2%) |  |
| **Race/ethnicity** |  |  |  |  |  | 0.32 |
| White | 156 (62.2%) | 2834 (65.6%) | 483 (70.0%) | 1813 (60.6%) | 5286 (64.1%) |  |
| Black | 39 (15.5%) | 555 (12.9%) | 100 (14.5%) | 381 (12.7%) | 1075 (13.0%) |  |
| Hispanic | 31 (12.4%) | 514 (11.9%) | 74 (10.7%) | 412 (13.8%) | 1031 (12.5%) |  |
| Asian | 11 (4.4%) | 250 (5.8%) | 20 (2.9%) | 250 (8.4%) | 531 (6.4%) |  |
| Other, unknown, missing | 14 (5.6%) | 165 (3.8%) | 13 (1.9%) | 137 (4.6%) | 329 (4.0%) |  |
| **Annual household income** |  |  |  |  |  | 0.46 |
| <$40,000 | 74 (29.5%) | 1090 (25.2%) | 125 (18.1%) | 663 (22.2%) | 1952 (23.7%) |  |
| $40,000 - $74,999 | 55 (21.9%) | 1231 (28.5%) | 166 (24.1%) | 780 (26.1%) | 2232 (27.0%) |  |
| $75,000 – $124,999 | 73 (29.1%) | 1177 (27.3%) | 232 (33.6%) | 786 (26.3%) | 2268 (27.5%) |  |
| $125,000 – $199,999 | >11* | 437 (10.1%) | >100* | 416 (13.9%) | 971 (11.8%) |  |
| ≥200,000 | <11* | 159 (3.7%) | >40* | 186 (6.2%) | 396 (4.8%) |  |
| Unknown/missing | 24 (9.6%) | 224 (5.2%) | 23 (3.3%) | 162 (5.4%) | 433 (5.2%) |  |
| **Baseline Hb_A1c_** |  |  |  |  |  | 0.28 |
| Mean (SD) | 7.6 (0.5) | 7.7 (0.5) | 7.5 (0.5) | 7.6 (0.5) | 7.6 (0.5) |  |
| Median (IQR) | 7.7 (7.2, 8.0) | 7.7 (7.3, 8.1) | 7.5 (7.1, 7.9) | 7.5 (7.2, 8.0) | 7.6 (7.2, 8.0) |  |
| **Baseline HbA_1c_ categories** |  |  |  |  |  | 0.29 |
| 6.8-6.9% | 27 (10.8%) | 295 (6.8%) | 100 (14.5%) | 289 (9.7%) | 711 (8.6%) |  |
| 7.0-7.9% | 145 (57.8%) | 2651 (61.4%) | 431 (62.5%) | 1943 (64.9%) | 5170 (62.7%) |  |
| 8.0-8.5% | 79 (31.5%) | 1372 (31.8%) | 159 (23.0%) | 761 (25.4%) | 2371 (28.7%) |  |
| **Baseline creatinine** |  |  |  |  |  | 0.25 |
| N (%) | 167 (66.5%) | 2905 (67.3%) | 502 (72.8%) | 2248 (75.1%) | 5822 (70.6%) |  |
| Mean (SD) | 0.9 (0.2) | 0.9 (0.2) | 0.8 (0.2) | 0.9 (0.2) | 0.9 (0.2) |  |
| Median (IQR) | 0.9 (0.7, 1.0) | 0.9 (0.7, 1.0) | 0.8 (0.7, 1.0) | 0.9 (0.7, 1.0) | 0.9 (0.7, 1.0) |  |
| **Baseline comorbidities** |  |  |  |  |  |  |
| Nephropathy | 41 (16.3%) | 412 (9.5%) | 41 (5.9%) | 243 (8.1%) | 737 (8.9%) | 0.33 |
| Retinopathy | 22 (8.8%) | 205 (4.7%) | 29 (4.2%) | 166 (5.5%) | 422 (5.1%) | 0.19 |
| Neuropathy | 55 (21.9%) | 561 (13.0%) | 84 (12.2%) | 325 (10.9%) | 1025 (12.4%) | 0.30 |
| Hyperglycemia | <11* | <11* | 0 (0.0%) | 0 (0.0%) | <11* | 0.18 |
| Hypoglycemia | <11* | <11* | 0 (0.0%) | 0 (0.0%) | <11* | 0.09 |
| Coronary artery disease | 26 (10.4%) | 395 (9.1%) | 38 (5.5%) | 247 (8.3%) | 706 (8.6%) | 0.18 |
| Chronic kidney disease | 21 (8.4%) | 141 (3.3%) | 11 (1.6%) | 92 (3.1%) | 265 (3.2%) | 0.32 |
| Cerebrovascular disease | <11* | 126 (2.9%) | ≥11* | 97 (3.2%) | 246 (3.0%) | 0.07 |
| Peripheral vascular disease | 21 (8.4%) | 217 (5.0%) | 21 (3.0%) | 141 (4.7%) | 400 (4.8%) | 0.23 |
| **Baseline medications** |  |  |  |  |  |  |
| Statin | 145 (57.8%) | 2976 (68.9%) | 415 (60.1%) | 2092 (69.9%) | 5628 (68.2%) | 0.25 |
| Non-statin lipid lower medications | 31 (12.4%) | 516 (11.9%) | 99 (14.3%) | 464 (15.5%) | 1110 (13.5%) | 0.09 |
| ACE inhibitor | 119 (47.4%) | 1958 (45.3%) | 279 (40.4%) | 1225 (40.9%) | 3581 (43.4%) | 0.14 |
| Angiotensin receptor blocker | 44 (17.5%) | 1121 (26.0%) | 201 (29.1%) | 829 (27.7%) | 2195 (26.6%) | 0.28 |
| ACE inhibitor or angiotensin receptor blocker | 157 (62.5%) | 2994 (69.3%) | 465 (67.4%) | 1997 (66.7%) | 5613 (68.0%) | 0.14 |
| Sacubitril /Valsartan | 0 (0.0%) | 0 (0.0%) | 0 (0.0%) | 0 (0.0%) | 0 (0.0%) | -- |
| Warfarin | <11* | 93 (2.2%) | ≥11* | 45 (1.5%) | 160 (1.9%) | 0.06 |
| Direct oral anti-coagulant | <11* | 56 (1.3%) | ≥11* | 51 (1.7%) | 128 (1.6%) | 0.08 |
| Peripheral neuropathy medications | 46 (18.3%) | 421 (9.7%) | 79 (11.4%) | 304 (10.2%) | 850 (10.3%) | 0.25 |
| **Specialty of treating physicians** |  |  |  |  |  | 0.45 |
| Primary care | 153 (61.0%) | 3429 (79.4%) | 450 (65.2%) | 2204 (73.6%) | 6236 (75.6%) |  |
| Endocrinology | 28 (11.2%) | 148 (3.4%) | 75 (10.9%) | 140 (4.7%) | 391 (4.7%) |  |
| Cardiology | <11* | >20* | <11* | ≥20* | 58 (0.7%) |  |
| Nephrology | <11* | <11* | <11* | <11* | 15 (0.2%) |  |
| Other | 20 (8.0%) | 246 (5.7%) | 102 (14.8%) | 192 (6.4%) | 560 (6.8%) |  |
| Unknown | 49 (19.5%) | 460 (10.7%) | 59 (8.6%) | 424 (14.2%) | 992 (12.0%) |  |
| **Year of cohort entry** |  |  |  |  |  | 0.27 |
| 2010 | <11* | 236 (5.5%) | ≥30* | 191 (6.4%) | 477 (5.8%) |  |
| 2011 | 14 (5.6%) | 216 (5.0%) | 37 (5.4%) | 224 (7.5%) | 491 (6.0%) |  |
| 2012 | 19 (7.6%) | 275 (6.4%) | 40 (5.8%) | 253 (8.5%) | 587 (7.1%) |  |
| 2013 | 23 (9.2%) | 357 (8.3%) | 73 (10.6%) | 319 (10.7%) | 772 (9.4%) |  |
| 2014 | 21 (8.4%) | 423 (9.8%) | 71 (10.3%) | 397 (13.3%) | 912 (11.1%) |  |
| 2015 | 22 (8.8%) | 514 (11.9%) | 75 (10.9%) | 252 (8.4%) | 863 (10.5%) |  |
| 2016 | 41 (16.3%) | 635 (14.7%) | 116 (16.8%) | 343 (11.5%) | 1135 (13.8%) |  |
| 2017 | 42 (16.7%) | 861 (19.9%) | 119 (17.2%) | 471 (15.7%) | 1493 (18.1%) |  |
| 2018 | 49 (19.5%) | 670 (15.5%) | 96 (13.9%) | 446 (14.9%) | 1261 (15.3%) |  |
| 2019 | <11* | 131 (3.0%) | ≥20* | 97 (3.2%) | 261 (3.2%) |  |

Abbreviations: SMD, standardized mean difference

* Cell suppression based on OptumLabs Cell Size Suppression Rules. N<11 are masked to protect patient confidentiality

Table S10. Pairwise standardized mean difference for weighted cohort.

|  | **Glimepiride vs Liraglutide** | **Glimepiride vs Sitagliptin** | **Liraglutide vs Sitagliptin** |
| --- | --- | --- | --- |
| **Age** | 0.14 | 0.00 | 0.14 |
| **Sex** | 0.05 | 0.02 | 0.03 |
| **Race/ethnicity** | 0.08 | 0.02 | 0.09 |
| **Annual Household Income** | 0.10 | 0.03 | 0.10 |
| **Baseline HbA_1c_** | 0.06 | 0.03 | 0.03 |
| **Baseline HbA_1c_ categories** | 0.09 | 0.02 | 0.07 |
| **Baseline creatinine** | 0.02 | 0.00 | 0.02 |
| **Baseline comorbidities** |  |  |  |
| Nephropathy | 0.03 | 0.02 | 0.05 |
| Retinopathy | 0.03 | 0.01 | 0.03 |
| Neuropathy | 0.03 | 0.01 | 0.05 |
| Hyperglycemia | 0.03 | 0.03 | 0.00 |
| Hypoglycemia | 0.03 | 0.03 | 0.00 |
| Coronary artery disease | 0.02 | 0.02 | 0.04 |
| Chronic kidney disease | 0.01 | 0.00 | 0.01 |
| Cerebrovascular disease | 0.00 | 0.00 | 0.00 |
| Peripheral vascular disease | 0.02 | 0.00 | 0.01 |
| **Baseline medications** |  |  |  |
| ACE inhibitor | 0.00 | 0.01 | 0.01 |
| ARB | 0.02 | 0.01 | 0.01 |
| ACE inhibitor or ARB | 0.03 | 0.01 | 0.04 |
| Direct oral anticoagulant | 0.01 | 0.00 | 0.01 |
| Statin | 0.07 | 0.02 | 0.08 |
| Non-statin lipid lowering medications | 0.05 | 0.01 | 0.04 |
| Warfarin | 0.00 | 0.02 | 0.02 |
| Peripheral neuropathy medications | 0.09 | 0.01 | 0.09 |
| **Specialty of treating physicians** | 0.10 | 0.02 | 0.10 |
| **Year of cohort entry** | 0.13 | 0.03 | 0.13 |

Figure S4. Inverse probability of treatment weighted Kaplan Meier curve for the time to right censoring by treatment groups.

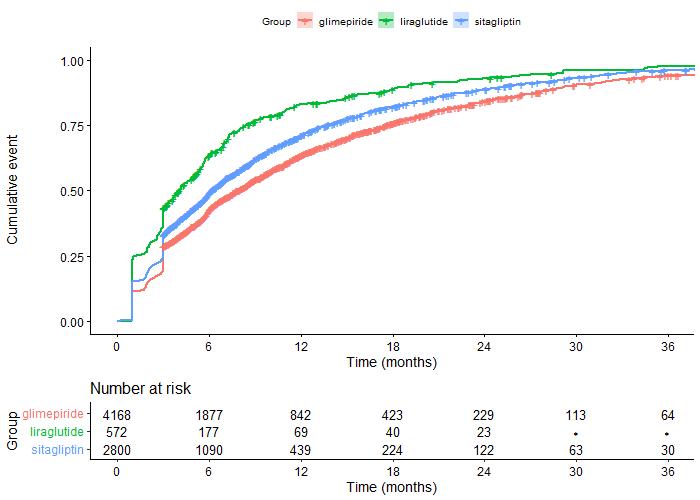
Figure S5. Cumulative risks of secondary metabolic failure in propensity score weighted patients. Censor per-protocol primary analysis.

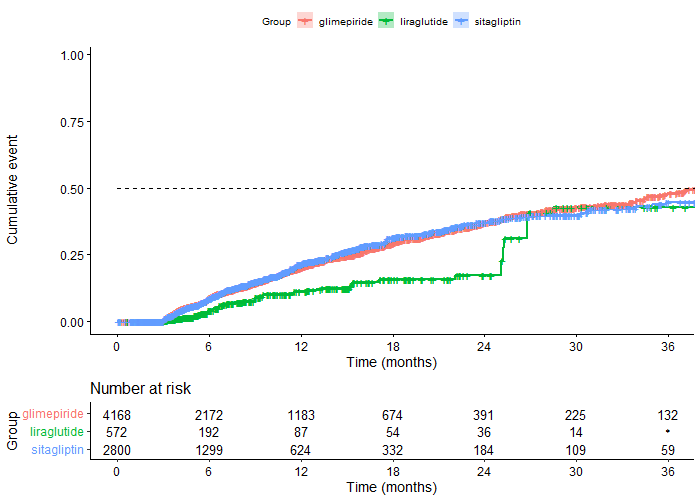

| **Time to secondary metabolic failure** | **First Quintile** | **Median** |
| --- | --- | --- |
| Glimepiride | 472 (95% CI, 415-498) | 1237 (95% CI, 1054-1478) |
| Liraglutide | 768 (95% CI, 764-NA) | 1601 (95% CI, 815-NA) |
| Sitagliptin | 437 (95% CI, 383-483) | 1521 (95% CI, 1155-NA) |

Table S11. Rates of secondary microvascular, macrovascular, and safety endpoints (weighted)**.** Rates of end-stage kidney disease, heart failure, pancreatitis, pancreatic/thyroid cancer, and all-cause mortality are not shown because there were fewer than 11 crude (unweighted) events in at least one of the treatment arms.

|  | **No. events** | **Event rate per 100 person-years** |
| --- | --- | --- |
| **Retinopathy** |  |  |
| Glimepiride | 188 | 1.83 |
| Liraglutide | 31 | 2.16 |
| Sitagliptin | 119 | 1.70 |
| Total | 338 | 1.80 |
| **Neuropathy** |  |  |
| Glimepiride | 609 | 5.92 |
| Liraglutide | 97 | 6.76 |
| Sitagliptin | 364 | 5.19 |
| Total | 1070 | 5.70 |
| **MACE** |  |  |
| Glimepiride | 158 | 1.54 |
| Liraglutide | 25 | 1.74 |
| Sitagliptin | 96 | 1.37 |
| Total | 279 | 1.49 |
| **Other Cardiovascular events** |  |  |
| Glimepiride | 204 | 1.98 |
| Liraglutide | 33 | 2.30 |
| Sitagliptin | 122 | 1.74 |
| Total | 359 | 1.91 |
| **Cancer** |  |  |
| Glimepiride | 175 | 1.70 |
| Liraglutide | 27 | 1.88 |
| Sitagliptin | 124 | 1.77 |
| Total | 326 | 1.74 |
| **Any hospital admission** |  |  |
| Glimepiride | 734 | 7.13 |
| Liraglutide | 102 | 7.11 |
| Sitagliptin | 480 | 6.84 |
| Total | 1316 | 7.02 |

Table S12. Hazard ratios of secondary microvascular, macrovascular, and safety endpoints**.**

|  | **HR (95% CI)** | **Holm adjusted p-value** |
| --- | --- | --- |
| **Retinopathy** |  |  |
| Liraglutide vs Glimepiride | 1.22 (0.75, 1.96) | 0.84 |
| Sitagliptin vs Glimepiride | 0.93 (0.73, 1.17) | 0.84 |
| Liraglutide vs Sitagliptin | 1.31 (0.80, 2.14) | 0.83 |
| **Neuropathy** |  |  |
| Liraglutide vs Glimepiride | 1.18 (0.90, 1.55) | 0.23 |
| Sitagliptin vs Glimepiride | 0.87 (0.76, 0.99) | 0.09 |
| Liraglutide vs Sitagliptin | 1.36 (1.03, 1.80) | 0.09 |
| **MACE** |  |  |
| Liraglutide vs Glimepiride | 1.13 (0.61, 2.10) | 1.00 |
| Sitagliptin vs Glimepiride | 0.89 (0.69, 1.15) | 1.00 |
| Liraglutide vs Sitagliptin | 1.27 (0.67, 2.40) | 1.00 |
| **Other Cardiovascular events** |  |  |
| Liraglutide vs Glimepiride | 1.17 (0.71, 1.94) | 0.70 |
| Sitagliptin vs Glimepiride | 0.87 (0.69, 1.09) | 0.70 |
| Liraglutide vs Sitagliptin | 1.35 (0.80, 2.27) | 0.70 |
| **Cancer** |  |  |
| Liraglutide vs Glimepiride | 1.11 (0.65, 1.89) | 1.00 |
| Sitagliptin vs Glimepiride | 1.04 (0.83, 1.31) | 1.00 |
| Liraglutide vs Sitagliptin | 1.06 (0.62, 1.83) | 1.00 |
| **Any hospital admission** |  |  |
| Liraglutide vs Glimepiride | 1.02 (0.78, 1.33) | 1.00 |
| Sitagliptin vs Glimepiride | 0.95 (0.85, 1.07) | 1.00 |
| Liraglutide vs Sitagliptin | 1.07 (0.81, 1.40) | 1.00 |

Table S13. Subgroup Analyses for Primary Metabolic Failure**.**

|  | **Hazard ratio (95% CI)** | **Holm adjusted p-value** |
| --- | --- | --- |
| **Baseline HbA_1c_ level** |  |  |
| **Baseline HbA_1c_ < 7.0%** |  |  |
| Liraglutide vs Glimepiride | 0.21 (0.06, 0.79) | 0.06 |
| Sitagliptin vs Glimepiride | 0.93 (0.61, 1.43) | 0.75 |
| Liraglutide vs Sitagliptin | 0.23 (0.06, 0.85) | 0.06 |
| **Baseline HbA_1c_ ≥ 7.0%** |  |  |
| Liraglutide vs Glimepiride | 0.59 (0.44, 0.78) | <0.001 |
| Sitagliptin vs Glimepiride | 1.04 (0.94, 1.14) | 0.44 |
| Liraglutide vs Sitagliptin | 0.58 (0.43, 0.79) | <0.001 |
| **Age group** |  |  |
| **Age < 65** |  |  |
| Liraglutide vs Glimepiride | 0.54 (0.42, 0.71) | <0.001 |
| Sitagliptin vs Glimepiride | 0.93 (0.82, 1.07) | 0.30 |
| Liraglutide vs Sitagliptin | 0.58 (0.44, 0.77) | <0.001 |
| **Age ≥ 65** |  |  |
| Liraglutide vs Glimepiride | 0.57 (0.31, 1.06) | 0.10 |
| Sitagliptin vs Glimepiride | 1.14 (1.00, 1.29) | 0.10 |
| Liraglutide vs Sitagliptin | 0.50 (0.27, 0.94) | 0.09 |
| **Gender** |  |  |
| **Female** |  |  |
| Liraglutide vs Glimepiride | 0.49 (0.33, 0.74) | 0.002 |
| Sitagliptin vs Glimepiride | 0.99 (0.87, 1.13) | 0.88 |
| Liraglutide vs Sitagliptin | 0.50 (0.33, 0.75) | 0.002 |
| **Male** |  |  |
| Liraglutide vs Glimepiride | 0.63 (0.42, 0.93) | 0.04 |
| Sitagliptin vs Glimepiride | 1.07 (0.94, 1.22) | 0.31 |
| Liraglutide vs Sitagliptin | 0.59 (0.39, 0.88) | 0.03 |
| **Race/Ethnicity** |  |  |
| **White** |  |  |
| Liraglutide vs Glimepiride | 0.58 (0.42, 0.81) | 0.002 |
| Sitagliptin vs Glimepiride | 1.06 (0.94, 1.19) | 0.34 |
| Liraglutide vs Sitagliptin | 0.55 (0.40, 0.77) | 0.001 |
| **Black** |  |  |
| Liraglutide vs Glimepiride | 0.57 (0.23, 1.41) | 0.65 |
| Sitagliptin vs Glimepiride | 0.84 (0.64, 1.11) | 0.65 |
| Liraglutide vs Sitagliptin | 0.68 (0.27, 1.70) | 0.65 |
| **Hispanic** |  |  |
| Liraglutide vs Glimepiride | 0.10 (0.03, 0.35) | <0.001 |
| Sitagliptin vs Glimepiride | 1.24 (0.95, 1.60) | 0.11 |
| Liraglutide vs Sitagliptin | 0.08 (0.03, 0.29) | <0.001 |
| **Asian** |  |  |
| Liraglutide vs Glimepiride | 0.84 (0.35, 2.03) | 1.00 |
| Sitagliptin vs Glimepiride | 0.70 (0.48, 1.01) | 0.17 |
| Liraglutide vs Sitagliptin | 1.20 (0.49, 2.94) | 1.00 |

Table S14. Subgroup Analyses for Secondary Metabolic Failure**.**

|  | **HR (95% CI)** | **Holm adjusted p-value** |
| --- | --- | --- |
| **Baseline HbA_1c_ level** |  |  |
| **Baseline HbA_1c_ < 7.0%** |  |  |
| Liraglutide vs Glimepiride | 0.28 (0.05, 1.55) | 0.43 |
| Sitagliptin vs Glimepiride | 0.81 (0.43, 1.54) | 0.53 |
| Liraglutide vs Sitagliptin | 0.34 (0.06, 1.92) | 0.45 |
| **Baseline HbA_1c_ ≥ 7.0%** |  |  |
| Liraglutide vs Glimepiride | 0.62 (0.43, 0.90) | 0.02 |
| Sitagliptin vs Glimepiride | 1.04 (0.91, 1.19) | 0.53 |
| Liraglutide vs Sitagliptin | 0.60 (0.41, 0.87) | 0.02 |
| **Age group** |  |  |
| **Age < 65** |  |  |
| Liraglutide vs Glimepiride | 0.53 (0.36, 0.78) | 0.004 |
| Sitagliptin vs Glimepiride | 0.98 (0.81, 1.18) | 0.80 |
| Liraglutide vs Sitagliptin | 0.54 (0.36, 0.81) | 0.005 |
| **Age ≥ 65** |  |  |
| Liraglutide vs Glimepiride | 0.68 (0.34, 1.38) | 0.62 |
| Sitagliptin vs Glimepiride | 1.08 (0.90, 1.30) | 0.62 |
| Liraglutide vs Sitagliptin | 0.63 (0.31, 1.29) | 0.62 |
| **Gender** |  |  |
| **Female** |  |  |
| Liraglutide vs Glimepiride | 0.57 (0.35, 0.91) | 0.06 |
| Sitagliptin vs Glimepiride | 0.99 (0.82, 1.20) | 0.94 |
| Liraglutide vs Sitagliptin | 0.57 (0.35, 0.93) | 0.06 |
| **Male** |  |  |
| Liraglutide vs Glimepiride | 0.65 (0.38, 1.12) | 0.24 |
| Sitagliptin vs Glimepiride | 1.08 (0.90, 1.29) | 0.41 |
| Liraglutide vs Sitagliptin | 0.60 (0.39, 0.91) | 0.22 |
| **Race/Ethnicity** |  |  |
| **White** |  |  |
| Liraglutide vs Glimepiride | 0.64 (0.42, 0.97) | 0.07 |
| Sitagliptin vs Glimepiride | 1.08 (0.92, 1.27) | 0.35 |
| Liraglutide vs Sitagliptin | 0.61 (0.39, 0.95) | 0.05 |
| **Black** |  |  |
| Liraglutide vs Glimepiride | 0.40 (0.15, 1.09) | 0.22 |
| Sitagliptin vs Glimepiride | 0.86 (0.59, 1.25) | 0.42 |
| Liraglutide vs Sitagliptin | 0.47 (0.17, 1.30) | 0.29 |
| **Hispanic** |  |  |
| Liraglutide vs Glimepiride | 0.13 (0.03, 0.63) | 0.03 |
| Sitagliptin vs Glimepiride | 0.97 (0.67, 1.41) | 0.88 |
| Liraglutide vs Sitagliptin | 0.14 (0.03, 0.65) | 0.03 |
| **Asian** |  |  |
| Liraglutide vs Glimepiride | 1.71 (0.89, 3.29) | 0.21 |
| Sitagliptin vs Glimepiride | 0.83 (0.47, 1.44) | 0.50 |
| Liraglutide vs Sitagliptin | 2.08 (1.05, 4.10) | 0.10 |

Figure S6. Cumulative risks of primary and secondary metabolic failure (sensitivity analysis). Sensitivity analysis conducted with as-treated censoring.

Primary metabolic failure

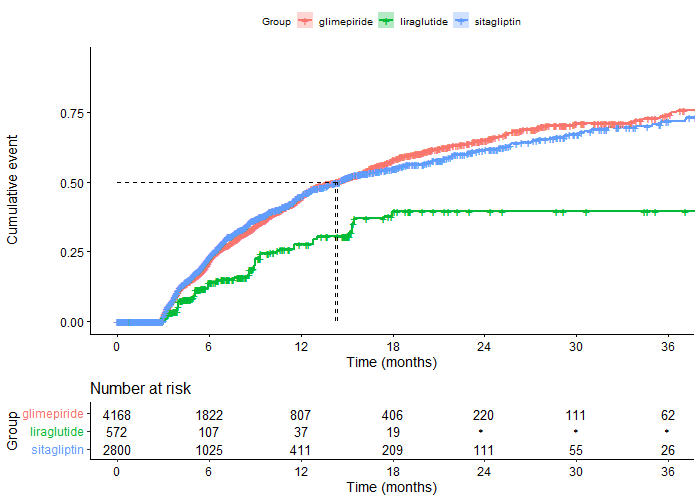

#### Secondary metabolic failure

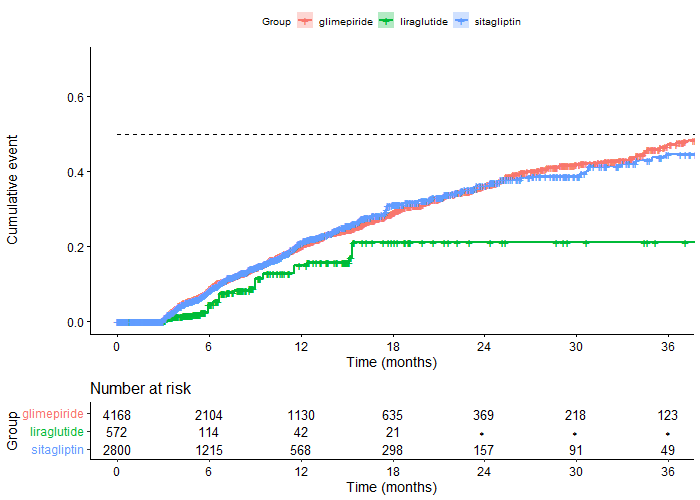
Table S15. Hazard ratios for primary and secondary metabolic failure (sensitivity analysis). Sensitivity analysis conducted with as-treated censoring.

|  | **HR (95% CI)** | **Holm adjusted p-value** |
| --- | --- | --- |
| **Primary metabolic failure** |  |  |
| Liraglutide vs Glimepiride | 0.43 (0.35, 0.80) | 0.006 |
| Sitagliptin vs Glimepiride | 1.03 (0.94, 1.13) | 0.56 |
| Liraglutide vs Sitagliptin | 0.51 (0.34, 0.78) | 0.006 |
| **Secondary metabolic failure** |  |  |
| Liraglutide vs Glimepiride | 0.59 (0.33, 1.07) | 0.20 |
| Sitagliptin vs Glimepiride | 1.03 (0.90, 1.19) | 0.62 |
| Liraglutide vs Sitagliptin | 0.57 (0.31, 1.04) | 0.20 |
| **Initiating insulin** |  |  |
| Liraglutide vs Glimepiride | 0.57 (0.20, 1.64) | 0.59 |
| Sitagliptin vs Glimepiride | 1.17 (0.77, 1.78) | 0.59 |
| Liraglutide vs Sitagliptin | 0.49 (0.17, 1.42) | 0.56 |

Table S16. Falsification end point analysis. Falsification endpoint was any office visit, emergency department, or hospital claim for pneumonia during the follow-up period.

|  | **HR (95% CI)** | **Holm adjusted p-value** |
| --- | --- | --- |
| Liraglutide vs Glimepiride | 1.01 (0.59, 1.73) | 1.00 |
| Sitagliptin vs Glimepiride | 0.97 (0.79, 1.21) | 1.00 |
| Liraglutide vs Sitagliptin | 1.04 (0.61, 1.80) | 1.00 |

Rationale: Our objective in choosing a falsification endpoint was to identify an outcome that is 1) measurable in our data, 2) is not related to any of the study outcomes, including glycemic control, microvascular complications, and macrovascular complications; and 3) is not related to glucose-lowering medications. Pneumonia meets each of these criteria. It also has a similar bias structure as HbA_1c_, as more clinically complex and/or sicker patients may be more likely to experience pneumonia and also to experience deterioration in glycemic control, have their HbA_1c_ tested more frequently, and experience microvascular and macrovascular complications.
